# Risk heterogeneity in compartmental HIV transmission models of ART as prevention in Sub-Saharan Africa: A scoping review

**DOI:** 10.1101/2021.03.29.21254586

**Authors:** Jesse Knight, Rupert Kaul, Sharmistha Mishra

## Abstract

**Background:** Transmission models provide complementary evidence to clinical trials about the potential population-level incidence reduction attributable to ART (ART prevention impact). Different modelling assumptions about risk heterogeneity may influence projected ART prevention impacts. We sought to review representations of risk heterogeneity in compartmental HIV transmission models applied to project ART prevention impacts in Sub-Saharan Africa.

**Methods:** We systematically reviewed studies published before January 2020 that used non-linear compartmental models of sexual HIV transmission to simulate ART prevention impacts in Sub-Saharan Africa. We summarized data on model structure/assumptions (factors) related to risk and intervention heterogeneity, and explored multivariate ecological associations of ART prevention impacts with modelled factors.

**Results:** Of 1384 search hits, 94 studies were included. 64 studies considered sexual activity stratification and 39 modelled at least one key population. 21 studies modelled faster/slower ART cascade transitions (HIV diagnosis, ART initiation, or cessation) by risk group, including 8 with faster and 4 with slower cascade transitions among key populations versus the wider population. In ecological analysis of 125 scenarios from 40 studies (subset without combination intervention), scenarios with risk heterogeneity that included turnover of higher risk groups were associated with smaller ART prevention benefits. Modelled differences in ART cascade across risk groups also influenced the projected ART benefits, including: ART prioritized to key populations was associated with larger ART prevention benefits. Of note, zero of these 125 scenarios considered lower ART coverage among key populations.

**Conclusion:** Among compartmental transmission models applied to project ART prevention impacts in Sub-Saharan Africa, representations of risk heterogeneity and projected impacts varied considerably. Inclusion/exclusion of risk heterogeneity with turnover, and intervention heterogeneity across risk groups could influence the projected impacts of ART scale-up. These findings highlight a need to capture risk heterogeneity with turnover and cascade heterogenetiy when projecting ART prevention impacts.

## 1 Introduction

As of 2019, two thirds (25.7 million) of all people living with HIV globally were in Sub-Saharan Africa (SSA), where an estimated one million new HIV infections were acquired in 2019 [1]. HIV treatment to reduce onward transmission remains a key element of combination HIV prevention [2]. Following empirical evidence of partnership-level efficacy of ART in preventing HIV transmission [3, 4, 5], and model-based evidence of “treatment as prevention” [6, 7, 8], several large-scale community-based trials of universal test-and-treat (UTT) were recently completed. These trials found that over 2-to-4 years, cumulative incidence under UTT did not significantly differ from cumulative incidence under ART according to national guidelines [9, 10, 11]. Thus the population-level reductions in incidence anticipated from transmission modelling were not observed in these trials [12, 13].

One theme in the proposed explanations for limited population-level ART prevention effectiveness was heterogeneity in intervention coverage and its intersection with heterogeneity in transmission risks [14, 12]. While viral suppression improved under UTT in all three trials, 21–54% of study participants remained unsuppressed [11, 9, 10]. Populations experiencing barriers to viral suppression under UTT may be at highest risk for acquisition and onward transmission, including key populations such as women and men engaged in sex work, and men who have sex with men [15, 16]. Data suggest subgroups not formally described as key populations, such as youth, and men who have sex with women, including clients of sex workers, may also experience barriers to engagement in ART care [17, 18, 19]. Indeed, data suggest UTT scale-up in practice has not reach subgroups equally [20].

Risk heterogeneity can be defined by various factors affecting acquisition and onward transmission risk, and is a well-established determinant of the emergence and persistence of HIV epidemics [21, 22]. Systematic model comparison studies by Hontelez et al. [23] and Rozhnova et al. [24] found that projected prevention impacts of ART scale-up were smaller when more heterogeneity was captured in the model. Given the upstream and complementary role of transmission modelling in estimating the prevention impacts of ART [7, 25], we sought to examine how heterogeneity in risk and ART uptake has been represented in mathematical models used to assess the prevention impacts of ART scale-up in SSA. We conducted a scoping review and ecological regression with the following objectives. Among non-linear compartmental models of sexual HIV transmission used to simulate the prevention impacts of ART in SSA:

1. In which epidemic contexts (geographies, populations, epidemic phases) have these models been applied?
2. How was the model structured to represent key factors of risk heterogeneity?
3. What are the potential influences of representations of risk heterogeneity on the projected prevention benefits of ART in the overall population?

## 2 Methods

We conducted a scoping review according to the PRISMA extension for scoping reviews (Appendix D).

### 2.1 Conceptual Framework for Risk Heterogeneity

We defined “factors of risk heterogeneity” as epidemiological phenomena and stratifications of populations, rates, or probabilities which may/not be included in transmission models. We defined 4 domains in which such factors might influence the transmission impact of ART:

- **Biological Effects**: differential transmission risk within HIV disease course that may coincide with differential ART coverage [26]
- **Behaviour Change Effects**: differential transmission risk due to behavioural changes related to engagement in the ART cascade [27, 28]
- **Network Effects**: differential transmission risk within sub-populations that increases the challenge of epidemic control through core group dynamics [22, 29, 30]
- **Cascade Effects**: differential transmission risk within sub-populations who experience barriers to ART care and achieving viral suppression, such as youth and key populations [31, 32, 15, 20]

We then compiled a list of key factors of risk heterogeneity, and their possible mechanisms of influence on ART prevention impact (Table 1).

**Table 1:** Factors of heterogeneity in HIV transmission and their possible mechanisms of influence on the prevention impact of ART interventions

| Factor | MP<sup>a</sup> | Definition | Possible mechanism(s) of influence on ART prevention impact |
| --- | --- | --- | --- |
| Acute Infection | $\beta_i$ | Increased infectiousness immediately following infection [33, 34] | **Biological:** transmissions during acute infection are unlikely to be prevented by ART |
| Late-Stage Infection | $\beta_i$ | Increased infectiousness during late-stage infection [33, 34] | **Biological:** transmissions during late-stage are more likely to be prevented by ART |
| Drug Resistance | $\beta_i$ | Transmitted factor that requires regimen switch to achieve viral suppression [35] | **Biological:** transmissions during longer delay to achieving viral suppression will not be prevented by ART |
| HIV Morbidity | $c; \eta$ | Reduced sexual activity during late-stage disease [36, 37] | **Behaviour Change:** reduced morbidity via ART could increase HIV prevalence among the sexually active population |
| HIV Counselling | $c; \eta; \kappa$ | Reduced sexual activity and/or increased condom use after HIV diagnosis [28] | **Behaviour Change:** increased HIV testing with ART scale up can contribute to prevention even before viral suppression is achieved |
| Activity Groups | $c; \kappa$ | Any stratification by rate of partnership formation [38] | **Network:** higher transmission risk among higher activity |
| Age Groups | $c; \kappa$ | Any stratification by age | **Network & Cascade:** higher transmission risk and barriers to viral suppression among youth [39, 20] |
| Key Populations | $c; \kappa$ | Any epidemiologically defined higher risk groups [40] | **Network & Cascade:** higher transmission risk and barriers to viral suppression among key populations [15] |
| Group Turnover | $\phi$ | Individuals move between activity groups and/or key populations reflecting sexual lifecourse [29] | **Network & Cascade:** counteract effect of stratification due to shorter periods in higher risk [41]; viral suppression may be achieved only after periods of higher risk |
| Assortative Mixing | $m$ | Any degree of assortative mixing (like-with-like) by age, activity, and/or key populations | **Network:** assortative sexual networks compound effect of stratification [38] |
| Partnership Types | $\eta; \kappa$ | Different partnership types are simulated, with different numbers of sex acts and/or condom usage [42] | **Network:** longer duration and lower condom use among main versus casual/sex work partnerships counteracts effect of stratification |
| ART Cascade Gaps | $\tau; \alpha$ | Slower ART cascade transitions among higher activity groups or key populations [15, 20] | **Cascade:** ART prevention benefits may be allocated differentially among risk groups |

<sup>a</sup> MP: Model Parameters — $\beta_i, \beta_s$ : transmission probability per act (infectiousness, susceptibility); $\eta$ : number of sex acts of each type per partnership; $\kappa$ : proportion of sex acts unprotected by a condom; $c$ : partnership formation rate; $m$ : mixing matrix (probability of partnership formation); $\mu$ : mortality rate; $v$ : entry rate; $\phi$ : internal turnover between activity groups; $\tau$ : testing rate; $\alpha$ : ART initiation rate (and retention-related factors).

### 2.2 Search

We searched MEDLINE and EMBASE via Ovid using search terms related to Sub-Saharan Africa (SSA), HIV, and transmission modelling (Table A.1). Search results were de-duplicated and screened by title and abstract in Covidence [43], followed by full-text screening using the criteria below. One reviewer (JK) conducted the search, screening, and data extraction.

#### 2.2.1 Inclusion/Exclusion Criteria

Table A.2 lists complete inclusion/exclusion criteria and related definitions. We included peer-reviewed, primary modelling studies that used non-linear compartmental models of sexual HIV transmission to project the prevention impacts of ART in any setting within SSA. We included studies published in English anytime before Jan 1, 2020, that simulated at least one scenario with increasing ART coverage, possibly alongside other interventions. The included studies formed Dataset A, used to complete objectives 1 and 2. A subset of Dataset A formed Dataset B, used to complete objective 3. Studies in Dataset B met three additional criteria: 1) examined scale-up of ART coverage alone (versus combination intervention); 2) examined ART intervention for the whole population (versus ART prioritized to subgroups); and 3) reported HIV incidence reduction and/or cumulative HIV infections averted relative to a base-case scenario reflecting status quo.

### 2.3 Data Extraction

Data extraction used the full text and all available supplementary material. Data were extracted per-study for objectives 1 and 2, and per-scenario for objective 3, possibly including multiple time horizons. Detailed variables definitions are given in Appendix B.

#### 2.3.1 Epidemic Context

For objective 1, we extracted data on geography, epidemic phase, and key populations explicitly considered in the model. We categorized studies by country, SSA region, and scale of the simulated population (city, sub-national, national, regional). We classified epidemic size at time of ART intervention using overall HIV prevalence (low: *<*1%, medium: 1-10%, high: *>*10%), and epidemic phase using overall HIV incidence trend (increasing, increasing-but-stabilizing, stable/equilibrium, decreasing-but-stabilizing, and decreasing).

We extracted whether any of the following key populations were modelled: female sex workers (FSW); male clients of FSW (Clients); men who have sex with men (MSM); transgender individuals; people who inject drugs (PWID); and prisoners. FSW were defined as any female activity group meeting 3 criteria: *<*5% of the female population; *<*1/3 the client population size; and having *>*50× the partners per year of the lowest sexually active female activity group [44, 42]. Clients were defined as any male activity group described as clients of FSW, and being *>*3× the FSW population size. We also extracted whether any groups in the model were described as MSM, transgender, PWID, or prisoners.

#### 2.3.2 Factors of Risk Heterogeneity

For objective 2, we examined if/how the factors of risk heterogeneity outlined in Table 1 were simulated in each study. We examined the number of *risk groups* defined by sex/gender and/or sexual activity, and any *turnover* of individuals between activity groups and/or key populations.

We classified how *partnership types* were defined: generic (all partnerships equal); based only on the activity groups involved; or overlapping, such that different partnership types could be formed between the same two activity groups. We extracted whether partnerships considered different numbers of sex acts and condom use, and whether models simulated any degree of assortative *mixing* by activity groups (preference for like-with-like) versus proportionate (random) mixing. The number of *age groups* was extracted, and whether *mixing* by age groups was proportionate, strictly assortative, or assortative with age differences. We extracted whether age conferred any transmission risk beyond mixing, such as different partnership formation rates.

Finally, we extracted whether rates of HIV diagnosis, ART initiation, and/or ART discontinuation differed across risk strata (sex/gender, activity, key populations, and/or age), and if so, how they differed.

#### 2.3.3 Prevention Impact of ART Scale-Up

For objective 3, we extracted the following data for each intervention scenario within Dataset B: the years that ART scale-up started (*t_0_*) and stopped (*t_f_*); the final overall ART coverage achieved and/or the final ART initiation rate (per person-year among PLHIV not yet in care); the criteria for ART initiation (e.g. CD4 count); and the relative reduction in transmission probability on ART. Then, we extracted the relative reduction in incidence and/or proportion of infections averted relative to the base-case scenario for available time horizons since *t_0_*.

We conducted an ecological analyses across all modelled scenarios to examine the relationship between factors of risk heterogenetiy and projected ART impacts, adjusting for other factors that could influence impacts. For each factor of risk heterogeneity, we compared projected ART impacts (incidence reduction/infections averted) across different factor levels (whether or not, and how the factor was modelled). We estimated the effect of each factor level on ART impacts using linear multivariate regression, with generalized estimating equations [45] to control for clustering due to multiple estimates per study/scenario. Time since *t_0_* was included as a covariate, and two variables were removed due to missingness. No variable selection was used to avoid biasing effect estimates [46]. We also plotted impacts versus time since *t_0_*, stratified by factor levels.

## 3 Results

The search yielded 1384 publications, of which 94 studies were included (Figure 1). These studies (Dataset A, Appendix A.3) applied non-linear compartmental modelling to simulate ART scale-up in SSA, of which 40 reported infections averted/incidence reduction due to population-wide ART scale-up without combination intervention, relative to a base-case reflecting status quo (Dataset B).

**Figure 1:**
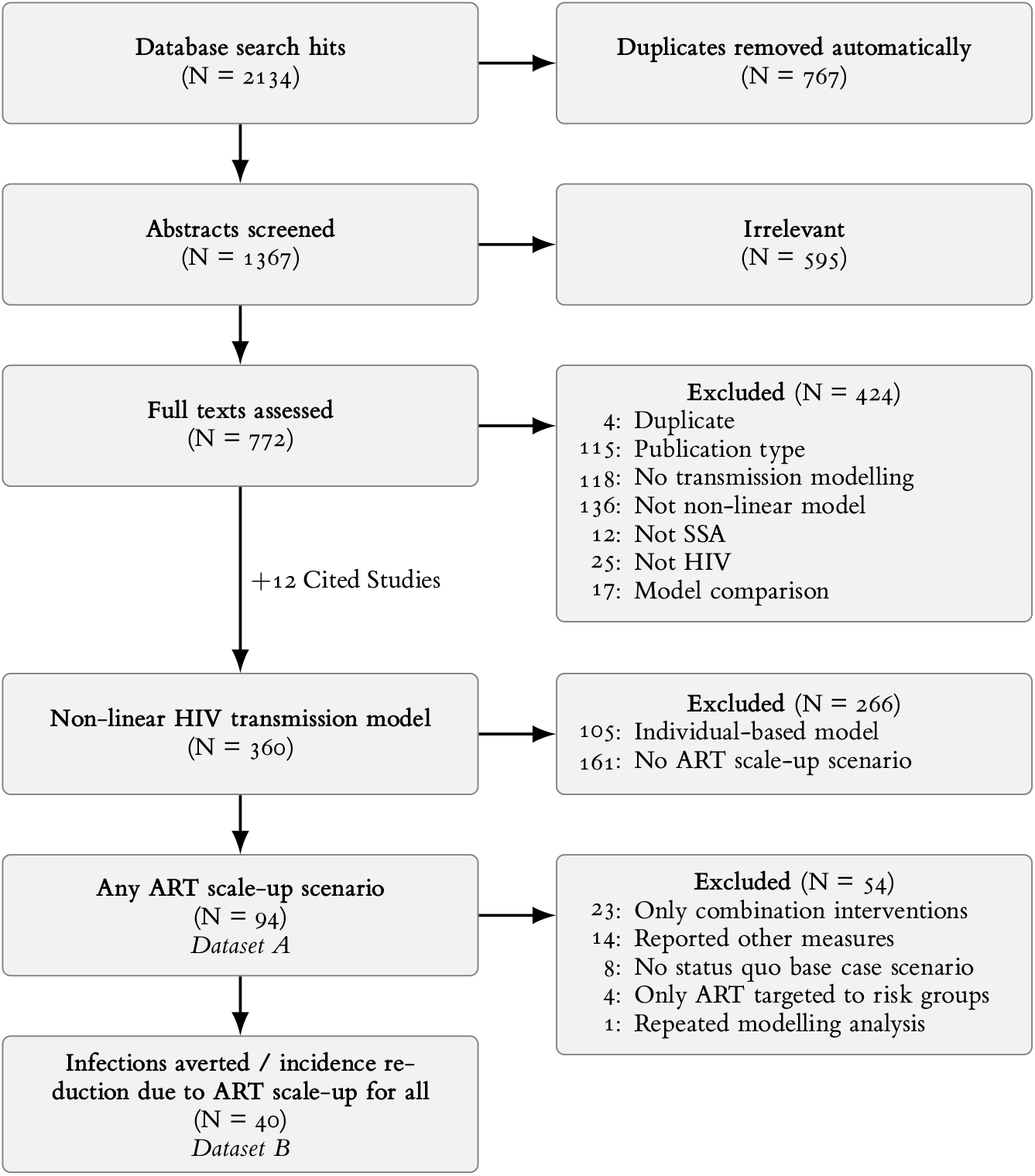
PRISMA flowchart of study identification

### 3.1 Epidemic Context

Table 2 summarizes key features of contexts within SSA where the prevention impacts of ART have been modelled. 61 studies modelled HIV transmission at the national level; studies also explored regional (1), subnational (16), and city-level (16) epidemic scales. South Africa was the most common country simulated (52 studies); Figure C.1 illustrates the number of studies by country. East Africa was the most represented SSA region, being simulated in 77 studies, followed by Southern (72), West (28), and Central Africa (13).

**Table 2:**
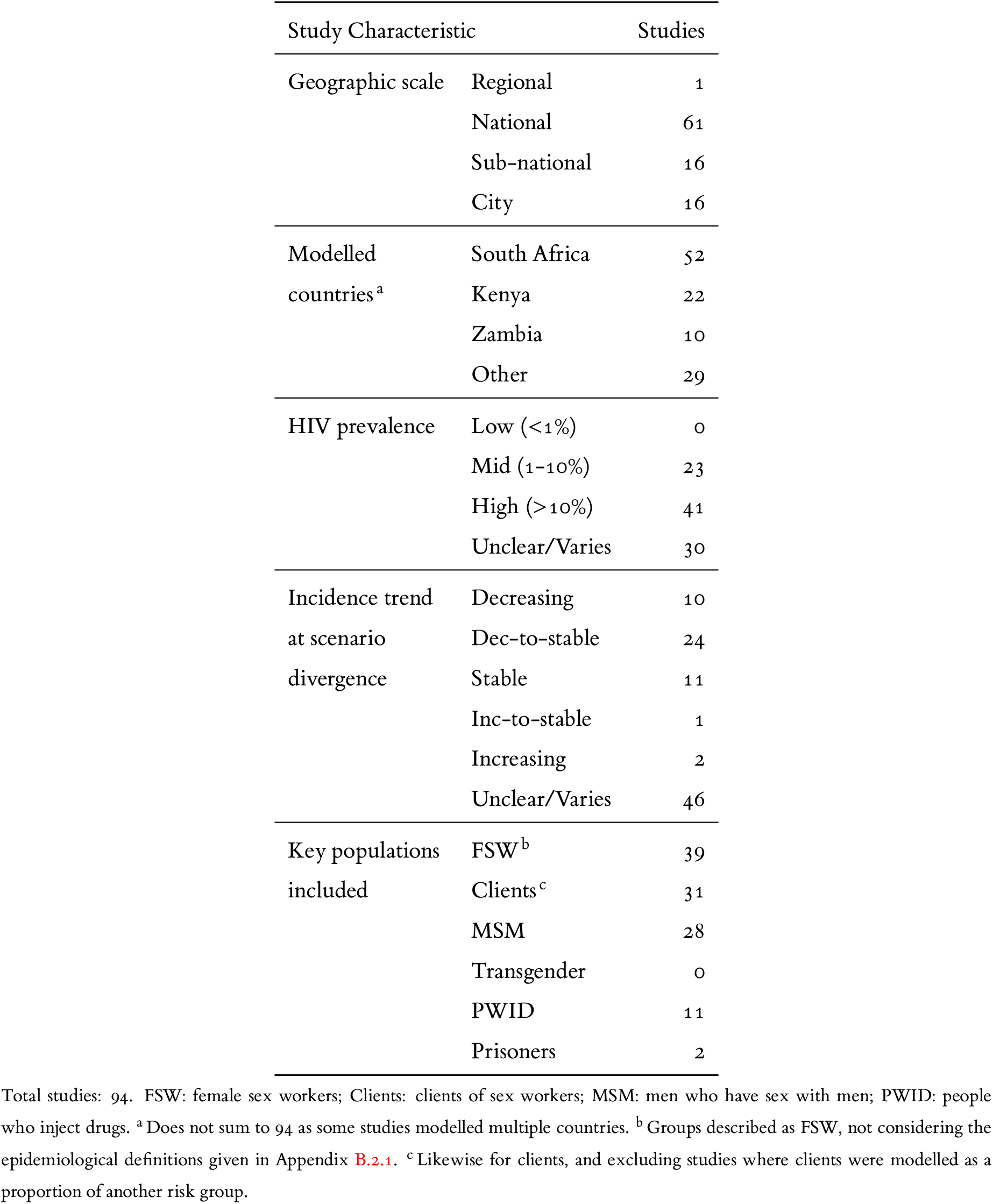
Summary of epidemic contexts within Sub-Saharan Africa where the prevention impacts of ART have been modelled

ART prevention impacts were most often modelled in high-prevalence (*>*10%) epidemics (41 studies) and medium-prevalence (1-10%) epidemics (23) (Figure C.2). No studies reported overall HIV prevalence of *<*1% at time of intervention, although for 30 studies, HIV prevalence was not reported or varied across simulated contexts/scenarios. The median [min, (IQR), max] year of intervention was 2014 [1990, (2010, 2015), 2040]; at which time HIV prevalence (%) was 15 [2, (6, 19), 32] (Figure C.2); and incidence (per 1000 person-years) was 14 [1, (9, 20), 50] (Figure C.3). Most reported incidence trends were decreasing or stable (45 of 48 reporting, Figure C.4).

#### 3.1.1 Key Populations

FSW were explicitly modelled in 39 studies. Among these (of studies where it was possible to evaluate): 21 (of 25) were *<*5% of the female population; 14 (of 24) were *<*1/3 the size of the client population; and 15 (of 22) had *>*50× partners per year versus the lowest sexually active female activity group. Clients of FSW were modelled as a unique group in 31 studies, among which 8 (of 17 reporting) were *>*3× the size of the FSW population. In another 8 studies, clients were defined as a proportion of another group, among which 6 (of 7) were *>*3× the FSW population size. Studies explicitly modelled men who have sex with men (MSM) in 28 studies; transgender in 0; people who inject drugs (PWID) in 11; and prisoners in 2.

### 3.2 Heterogeneity Factors

#### 3.2.1 Biological Effects

The median [min, (IQR), max] number of states used to represent HIV disease (ignoring treatment-related stratifications) was 5 [1, (3, 6), 25] (Figure C.5), and 2 studies represented HIV along a continuous dimension using partial differential equations. States of increased infectiousness associated with acute infection and late-stage disease were simulated in 68 and 74 studies, respectively.

The relative risk of HIV transmission on ART was 0.08 [0, (0.04, 0.13), 0.3] (Figure C.6), representing an average “on-treatment” state in 78 studies, versus a “virally suppressed” state in 15. Treatment failure due to drug resistance was simulated in 24 studies, including: 23 where individuals experiencing treatment failure were tracked separately from ART-naive; and 1 where such individuals transitioned back to a generic “off-treatment” state. Another 6 studies included a similar transition that was not identified as treatment failure versus ART cessation. Transmissible drug resistance was simulated in 9 studies.

#### 3.2.2 Behavioural Effects

Reduced sexual activity during late-stage HIV was simulated in 25 studies, including at least one state with: complete cessastion of sexual activity (14); reduced rate/number of partnerships (9); and/or reduced rate/number of sex acts per partnership (6).

Separate health states representing diagnosed HIV before treatment, and on-treatment before viral suppression were simulated in 30 and 17 studies, respectively. 22 studies modelled behaviour changes following awareness of HIV+ status, including: increased condom use (12); fewer partners per year (4); fewer sex acts per partnership (3); serosorting (1); and/or a generic reduction in transmission probability (8).

ART cessation was simulated in 35 studies, including: 16 where individuals no longer on ART were tracked separately from ART-naive; and 19 where such individuals transitioned back to a generic “off-treatment” state. Another 6 studies included a similar transition that was not identified as treatment failure versus ART cessation.

#### 3.2.3 Network Effects

Populations were stratified by activity (different rates and/or types of partnerships formed) in 59 studies, and by sex/gender in 64. The number of groups defined by sex/gender and/or activity was 6 [1, (2, 9), 19] (Figure C.7); and by activity alone (maximum number of groups among: women who have sex with men, men who have sex with women, MSM, or overall if sex/gender was not considered) was 3 [1, (1, 3), 18]. The highest activity groups for females and males (possibly including FSW/clients) comprised 2 [*<* 1, (2, 4), 23] and 9 [*<* 1, (2, 14), 35] % of female and male populations, respectively (Figures C.9 and C.10).

Turnover between activity groups and/or key populations was considered in 28 studies, of which 9 considered turnover of only one specific high-activity group or key population. Another 7 studies simulated movement only from lower to higher activity groups to re-balance group sizes against disproportionate HIV mortality.

Among 59 studies with activity groups, sexual mixing was modelled as assortative in 57 and proportionate in 2. Partnerships had equal probability of transmission in 39 studies, including all studies without activity groups. Partnerships were defined by the activity groups involved in 44 studies, among which transmission was usually lower in high-with-high activity partnerships than in low-with-low, due to fewer sex acts (31) and/or increased condom use (23). Transmission risk in high-with-low activity partnerships was defined by the: susceptible partner (9); lower activity partner (11); higher activity partner (3); or both partners’ activity groups (15); yielding indeterminate, higher, lower, or intermediate per-partnership transmission risk, respectively. Partnerships were defined based on overlapping types, such that different partnership types could be formed between the same two activity groups in 11 studies. All overlapping partnership types had differential total sex acts and condom use.

Age groups were simulated in 32 studies, among which, the number of age groups was 10 [2, (4, 34), 91] (Figure C.8), and 2 studies simulated age along a continuous dimension. Sexual mixing between age groups was assumed to be assortative either with (23) or without (3) average age differences between men and women; or proportionate (6). Differential risk behaviour by age was modelled in 29 studies.

#### 3.2.4 Cascade Effects

Differential transition rates along the ART cascade were considered in 21 studies, including differences between genders in 15; age groups in 7; and key populations in 12. Another 2 studies did not simulate differential cascade transitions, but justified the decision using context-specific data. Differences between genders included rates of HIV diagnosis (11); ART initiation (6); and ART cessation (1), with cascade engagement higher among women, in most cases attributed to antenatal services. Differences between age groups also affected rates of diagnosis (6); ART initiation (1); but not ART cessation (0). Among key populations, *lower* rates of diagnosis, ART initiation, and retention were simulated in 0, 2, and 4 studies respectively, while *higher* rates were simulated in 8, 2, and 1.

**Figure 2:**
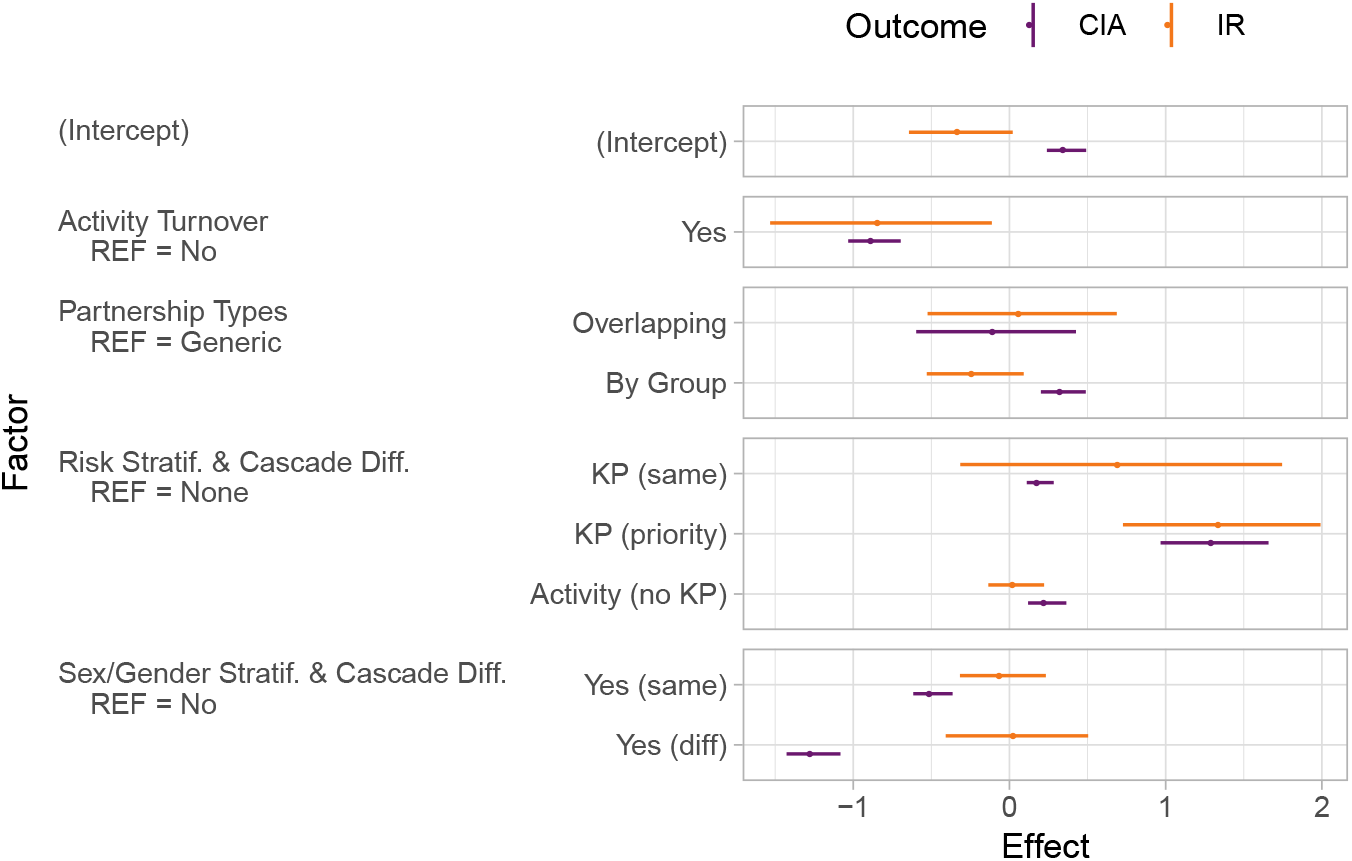
Effect estimates for selected factors of heterogeneity on incidence reduction (%, IR) and cumulative infections averted (%, CHI) from linear multivariate regression with generalized estimating equations. Numerical results given in Table 3. KP: key populations. priority: modelled ART cascade transitions were faster in KP vs overall due to prioritized programs; same: cascade transitions were assumed the same in KP as overall; diff: cascade transitions were slower among men. Factor definitions are given in Appendix B.

### 3.3 ART Prevention Impact

Dataset B comprised 40 studies, including 125 scenarios of ART scale-up. Relative incidence reduction (IR) with ART scale-up as compared to status quo was reported in 23 studies (61 scenarios); proportion of cumulative infections averted (CIA) due to ART scale-up in 24 (75); and 7 (11) reported both. Some scenarios included multiple time horizons.

Table 3 summarizes projected ART prevention impacts (IR, CIA), stratified by heterogeneity and contextual factors, plus adjusted effect estimates for each factor from multivariate analysis. Figures C.11–C.19 illustrate unadjusted impacts stratified by factor levels, while Figures C.20 and 2 (subset) illustrate effect estimates. Compared to models with homogeneous risk, including risk heterogeneity via static activity groups but without key populations was associated with slightly higher impacts—adjusted effect (95% CI): 4 (−14, 22)% IR, 24 (12, 36)% CIA. Including key population(s) and assuming similar ART cascade across groups was also associated with higher impact: 72 (−31, 175)% IR, 20 (11, 28)% CIA. However, including turnover of one/more higher risk group(s) was associated with smaller ART prevention impacts: −82 (−153, −11)% IR, −86 (−103, −70)% CIA. Taken together, models that captured heterogenetiy in risk across activity groups and/or key population(s) with turnover were associated with reduced ART prevention impacts.

**Table 3:**
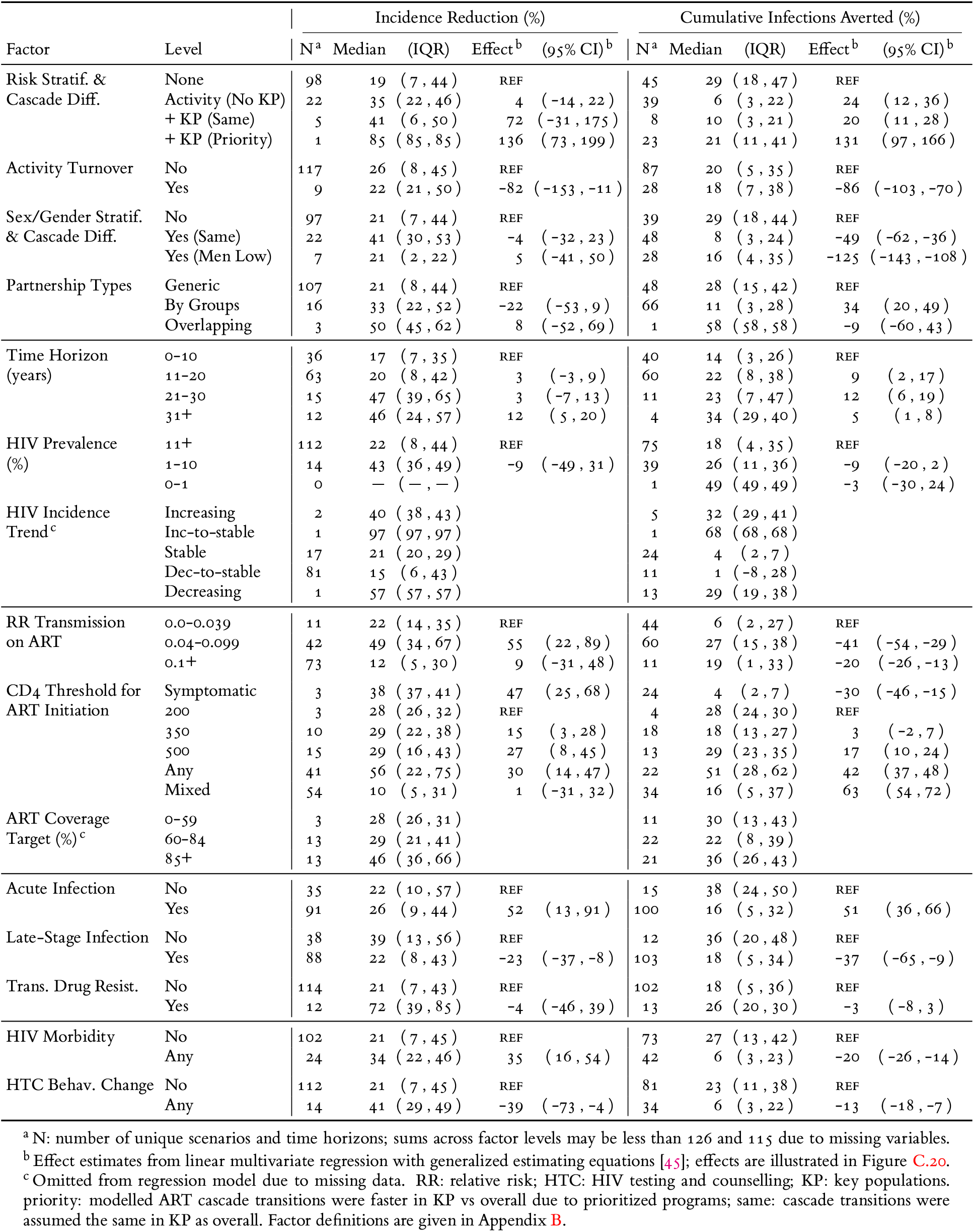
Projected ART prevention impacts, stratified by factors of risk heterogeneity and contexts

After including risk heterogeneity, further capturing differential ART cascade across activity groups or key populations was associated with differences in projected ART prevention impacts. Models stratified by sex/gender, and those that captured lower ART cascade among men were associated with a smaller CIA: −49 (−62, −36)% and −125 (−143, −108)%, respectively; although similar effects were not observed for IR: −4 (−32, 23)% and 5 (−41, 50)%. Where key populations were expliciltly modelled, including ART cascade prioritized to any key population(s) was associated with increased impact, enough to overcome reductions due to turnover: 136 (73, 199)% IR, 131 (97, 166)% CIA. No studies in Dataset B examined lower ART cascade among key population(s).

## 4 Discussion

Model-based evidence continues to support evaluation and mechanistic understanding of ART prevention impacts. Such evidence may be sensitive to modelling assumptions about risk heterogeneity. Via scoping review, we found that stratification by sexual activity and key population(s) was considered in approximately 2/3 and 2/5 of studies to date, respectively; 1/3 considered risk group turnover and 1/4 considered differential ART cascade by any risk group. In multivariate ecological analysis, we found that projected incidence reductions and propoportions of infections averted were influenced by risk heterogeneity when risk group turnover and differential ART cascade were also considered.

Our findings suggest that the proportion of onward transmission prevented through ART may be reduced via turnover. Data suggest considerable within-person variability in sexual risk among key populations, including MSM, FSW, and clients of FSW [47, 48, 49], as well as in the wider population [50]. This risk variability is often reflected in compartmental models as risk group turnover. Previous modelling suggested that turnover could *increase* the prevention benefits of treatment [51]; however, the model in [51] was calibrated to overall equilibrium prevalence, allowing the reproduction number to decrease with increasing turnover. By contrast, when calibrating to group-specific prevalence with turnover, greater risk heterogeneity is inferred with versus without turnover, and the reproduction number may actually increase [41]. Turnover of higher risk groups can also reduce ART coverage in those groups through net outflow of treated individuals, and net inflow of susceptible individuals, some of whom then become infected [41]. Thus, mechanistically, turnover could reduce the transmission benefits of ART. These findings suggest that turnover is important to capture as part of modeling risk heterogeneity, and as such, models would benefit from surveys, cohorts, and repeated population size estimates that can provide data on individual-level trajectories of sexual risk, such as duration in sex work [29].

Most models assumed equal ART cascade transition rates across subgroups, including diagnosis, ART initiation, and retention. However, recent data suggest differential ART cascade by sex, age, and key populations [32, 52, 53, 20]. These differences may stem from the unique needs of subgroups and is one reason why differentiated ART services are a core component of HIV programs [18, 54]. Moreover, barriers to ART may intersect with transmission risk, particularly among key populations, due to issues of stigma, discrimination, and criminalization [55, 12]. Our ecological analysis estimated that differences in cascade by sex (lower among men) or risk (key populations prioritized) had a large influence on projected ART prevention benefits. Thus, opportunities exist to incorporate differentiated cascade data, examine the intersections of intervention and risk heterogeneity, and to consider the impact of HIV services as delivered on the ground. Similar opportunities were noted regarding modelling of pre-exposure prophylaxis in SSA [56]. Depending on the research question, the modelled treatment cascade may need to include more cascade steps and states related to treatment failure/discontinuation.

The next generation of ART prevention impact modelling can be advanced by leveraging rapid growth in data on risk heterogeneity and its intersection with intervention heterogeneity [57, 58, 59]. Key populations often reflect intersections of risk heterogeneity with turnover, and intervention heterogeneity (cascade differences), which together suggest the unmet needs of key populations play an important role in the overall dynamics of HIV transmission in SSA [60, 61]. Although none of the models in the review considered a lower ART cascade among key populations, data suggest large cascade differences, most notably lower proportions across the cascade, among key populations in SSA [31, 15, 52]. Similarly, we found that the number of modelled clients per female sex worker, and the relative rate of partnership formation among female sex workers versus other women did not always reflect available data syntheses for sex work [29, 42]. Among studies with different partnership types, only 1/5 modelled main/spousal partnerships—with more sex acts/lower condom use— between two higher risk individuals, while 4/5 modelled only casual/commercial partnerships among higher risk individuals. However, data suggest that female sex workers form main/spousal partnerships with regular clients and boyfriends/spouses from higher risk groups [42]. Improved modelling and prioritization of sevices designed to reach key populations will rely on continued investment in community-led data collection for hard-to-reach populations.

Our scoping review has several limitations. First, we examined key populations based as traditionally defined [40], based on social and economic marginalization and criminalization in SSA, and future work would benefit from examing risk heterogenetiy across more subgroups, such as mobile populations and adolescent girls and young women, where data suggest cascade disparities and risk heterogeneity [62, 63]. Second, our conceptual framework for risk heterogeneity did not explicitly examine heterogeneity related to anal sex, which is associated with higher probability of HIV transmission; nor did we examine structural risk factors like violence [64, 65]. Third, we did not extract whether models were calibrated, and if so, which parameters were fixed versus fitted. If certain parameters were fitted, it could explain some counterintuitive effect estimates. For example, modelling increased infectiousness in late-stage HIV was associated with reduced ART prevention impacts. However, in most studies, newly ART-eligible patients via scale-up had earlier stage HIV; therefore, such patients would have lower modelled infectiousness than late-stage HIV, and lower infectiousness than in a model with uniform infectiousness fitted to the same data. A similar mechanism could explain increased ART prevention impacts when including acute infection. Importantly, we conducted an ecological analysis, and within-model comparisons like [30, 23] that explore the influence of each key factor identified in this review would be an important next step.

In conclusion, model-based evidence of ART prevention impacts could likely be improved by: 1) capturinig risk heterogeneity with risk group turnover, as a determinant of inferred risk heterogeneity during model calibration, and to reflect challenges to maintaining ART coverage among risk groups with high turnover; 2) integrating data on differences in ART cascade between sexual risk groups, to reflect services as delivered on the ground; and 3) capturing heterogenetiy in risks related to key populations, to reflect intersections of transmission risk and barriers to HIV services that may undermine the prevention benefits of ART.

## Data Availability

The extracted data and analysis code are available online via GitHub at the link below.

https://github.com/mishra-lab/sr-heterogeneity-hiv-models/

## Acknowledgements

We thank: Kristy Yiu (Unity Health Toronto) for research coordination support; Linwei Wang (Unity Health Toronto) for feedback on elements of the paper; and Kaitlin Fuller (University of Toronto) for her help designing the search strategy.

## Contributions

JK and SM conceptualized and designed the study, and developed the search strategy. JK performed the search, extracted the data, conducted the analysis, and generated the results. JK and SM drafted the manuscript and appendix. All authors (JK, RK, and SM) reviewed the results and contributed to writing the manuscript.

^1^WebPlotDigitizer: https://apps.automeris.io/wpd/

## appendix

## A Search Strategy

We designed our search strategy with guidance from an information specialist at our affiliate library (KF).

### A.1 Search Terms

Our search strategy and step-wise results are as follows, where term/ denotes a MeSH term, and .mp searches the main text fields, including title, abstract, and heading words. We searched MEDLINE and EMBASE via Ovid on 2020 March 20. Duplicate studies were removed automatically by Ovid and by Covidence; four additional duplicates with subtly different titles were later identified and removed manually.

**Table A.1:**
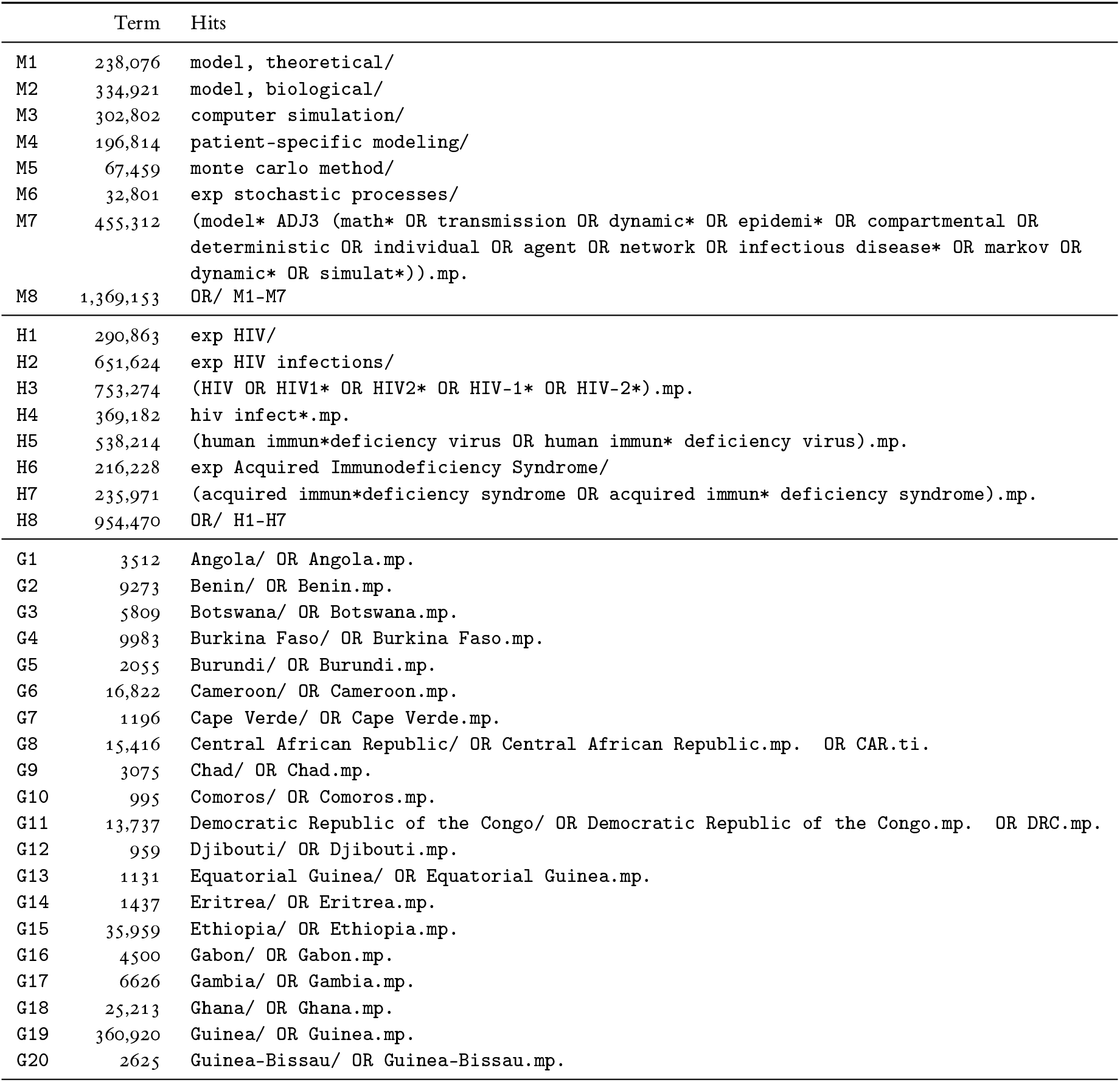
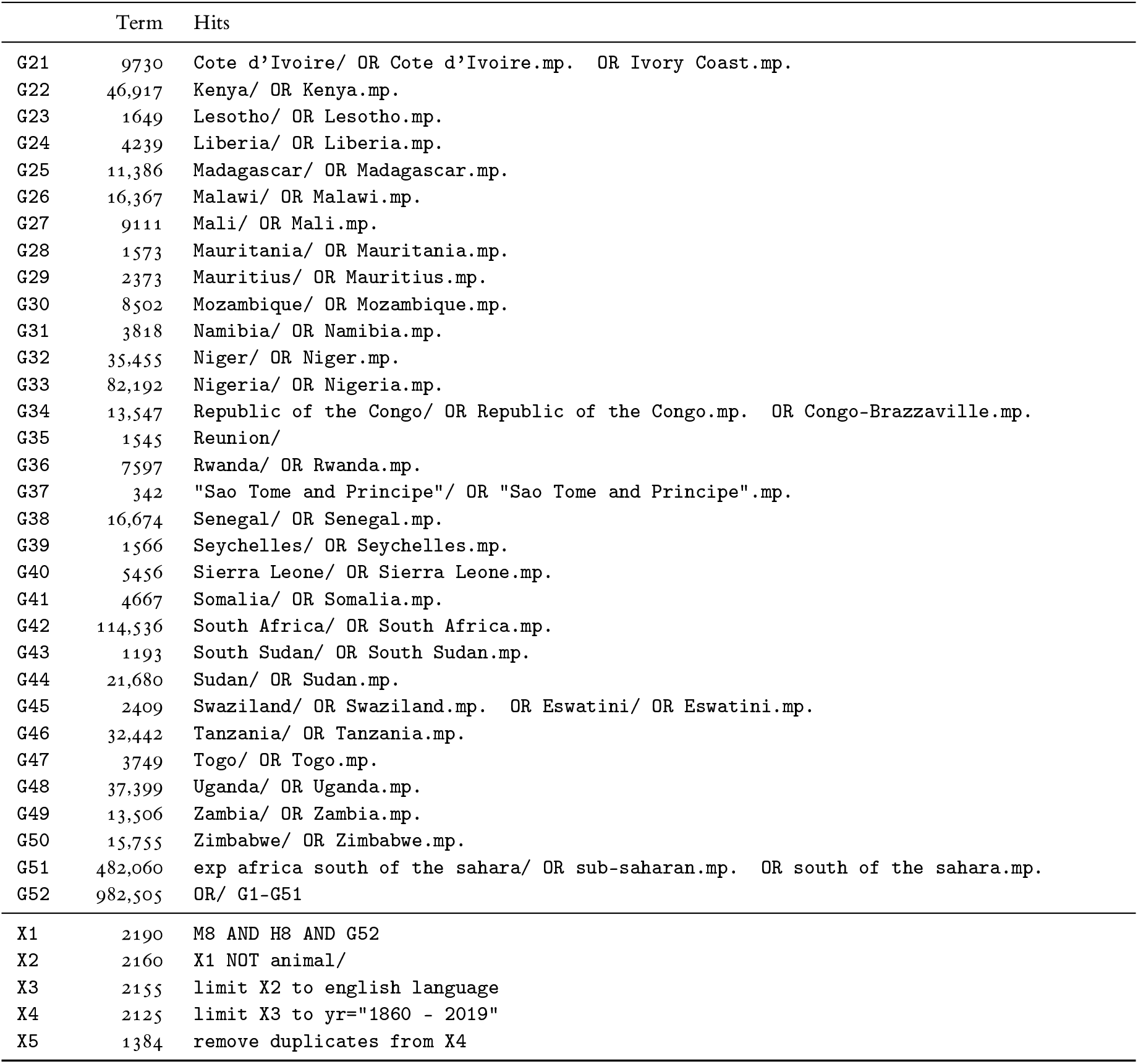
Search terms and hits

### A.2 Inclusion/Exclusion Criteria

**Table A.2:** Criteria for inclusion and exclusion

| Include | Exclude |
| --- | --- |
| **Publication Type** |  |
| English language<br>published before 2020<br>peer-reviewed journal article | non-English language<br>published in or after 2020<br>non-peer-reviewed article<br>review article<sup>1</sup><br>textbook, grey literature<br>opinions, comments, correspondence<br>conference abstracts and proceedings<br>model comparison study |
| **Mathematical Modelling of HIV Transmission** |  |
| sexual HIV transmission model<br>non-linear HIV transmission model<sup>2</sup><br>population-level dynamics<br>compartmental model<sup>3</sup> | no sexual HIV transmission modelled<br>HIV transmission model is linear<br>only within-host/cellular/protein modelling<br>individual-based model |
| **Context & Objectives** |  |
| any region in Sub-Saharan Africa (SSA)<sup>4</sup><br>assess prevention impact of ART scale-up for all<sup>5</sup> | only regions outside SSA modelled<br>only theoretical context modelled<br>only individual-level benefits of ART modelled<br>only prevention benefits of other interventions<br>no base-case scenario reflecting status quo<sup>*</sup><br>only ART-combination interventions<sup>*</sup><br>only ART intervention targeted to some risk groups<sup>*</sup><br>only ART prevention impacts reported for some risk groups<sup>*</sup><br>ART prevention impacts not reported<sup>5*</sup> |

<sup>1</sup> Review articles were included if they also presented new HIV transmission modelling results fitting our criteria. <sup>2</sup> We define a *non-linear model* as one where the number of infections projected at time $t$ is an iterative function of the number of infections previously projected by the model before time $t$ . <sup>3</sup> We define a *compartmental model* as one where the system variables represent the numbers of individuals in each state, rather than unique individuals. <sup>4</sup> SSA was defined based on the countries in the UN regions of East, South, Central, and West Africa, plus South Sudan (see Table A.1 for full country list). Studies were included if the model was parameterized/calibrated to reflect at least one context within SSA. Only model parameters & outcomes for SSA contexts were extracted. <sup>5</sup> Articles reporting HIV incidence reduction and/or cumulative HIV infections averted among the whole population due to increased coverage or initiation rate of ART for the whole population. <sup>\*</sup> Used to define Dataset B only.

### A.3 Included Studies

#### A.3.1 Dataset B

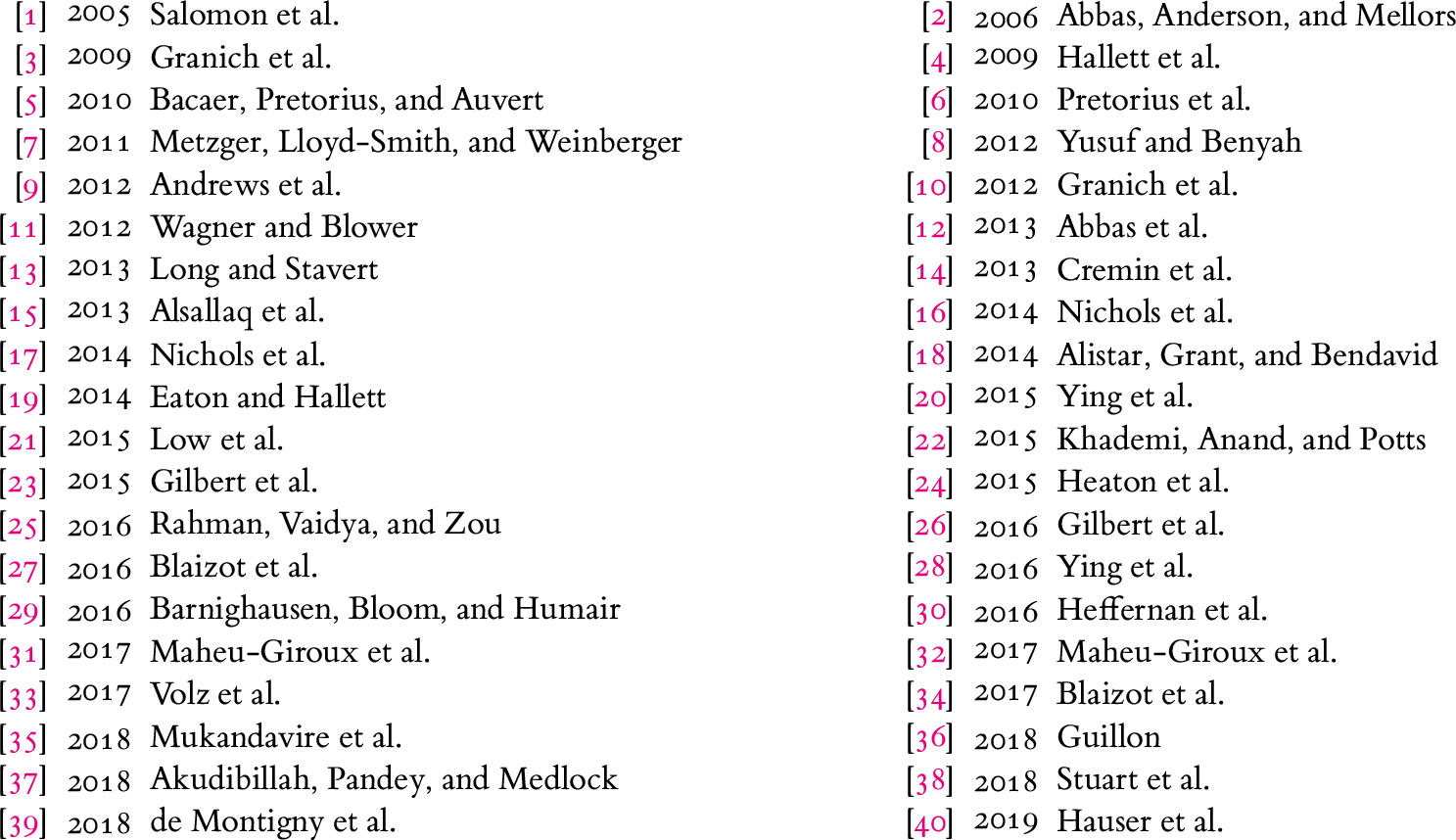

|  |  |
| --- | --- |
| [1] 2005 Salomon et al. | [2] 2006 Abbas, Anderson, and Mellors |
| [3] 2009 Granich et al. | [4] 2009 Hallett et al. |
| [5] 2010 Bacaer, Pretorius, and Auvert | [6] 2010 Pretorius et al. |
| [7] 2011 Metzger, Lloyd-Smith, and Weinberger | [8] 2012 Yusuf and Benyah |
| [9] 2012 Andrews et al. | [10] 2012 Granich et al. |
| [11] 2012 Wagner and Blower | [12] 2013 Abbas et al. |
| [13] 2013 Long and Stavert | [14] 2013 Cremin et al. |
| [15] 2013 Alsallaq et al. | [16] 2014 Nichols et al. |
| [17] 2014 Nichols et al. | [18] 2014 Alistar, Grant, and Bendavid |
| [19] 2014 Eaton and Hallett | [20] 2015 Ying et al. |
| [21] 2015 Low et al. | [22] 2015 Khademi, Anand, and Potts |
| [23] 2015 Gilbert et al. | [24] 2015 Heaton et al. |
| [25] 2016 Rahman, Vaidya, and Zou | [26] 2016 Gilbert et al. |
| [27] 2016 Blaizot et al. | [28] 2016 Ying et al. |
| [29] 2016 Barnighausen, Bloom, and Humair | [30] 2016 Heffernan et al. |
| [31] 2017 Maheu-Giroux et al. | [32] 2017 Maheu-Giroux et al. |
| [33] 2017 Volz et al. | [34] 2017 Blaizot et al. |
| [35] 2018 Mukandavire et al. | [36] 2018 Guillon |
| [37] 2018 Akudibillah, Pandey, and Medlock | [38] 2018 Stuart et al. |
| [39] 2018 de Montigny et al. | [40] 2019 Hauser et al. |

#### A.3.2 Dataset A less B

|  |  |
| --- | --- |
| [41] 2006 Johnson and Dorrington | [42] 2006 Baggaley, Garnett, and Ferguson |
| [43] 2006 Wilson, Kahn, and Blower | [44] 2008 Bacaer et al. |
| [45] 2009 Chigidi and Lungu | [46] 2010 Williams et al. |
| [47] 2011 Nyabadza and Mukandavire | [48] 2012 Barnighausen, Bloom, and Humair |
| [49] 2013 Wagner, Coburn, and Blower | [50] 2013 Decker et al. |
| [51] 2013 Wirtz et al. | [52] 2014 Shafer et al. |
| [53] 2014 Hove-Musekwa et al. | [54] 2014 Braithwaite et al. |
| [55] 2014 Nichols et al. | [56] 2014 Abu-Raddad and Awad |
| [57] 2014 Anderson et al. | [58] 2014 Alistar et al. |
| [59] 2014 Cori et al. | [60] 2014 Stover et al. |
| [61] 2014 Wirtz et al. | [62] 2015 Korenromp et al. |
| [63] 2015 Knight et al. | [64] 2015 Kerr et al. |
| [65] 2015 Fraser et al. | [66] 2015 Kassa and Ouhinou |
| [67] 2015 Bekker et al. | [68] 2015 Shannon et al. |
| [69] 2015 Blaizot et al. | [70] 2016 Smith et al. |
| [71] 2016 Atun et al. | [72] 2016 Shattock et al. |
| [73] 2016 McGillen et al. | [74] 2016 Johnson et al. |
| [75] 2016 Sharma et al. | [76] 2017 Akudibillah, Pandey, and Medlock |
| [77] 2017 Alsallaq et al. | [78] 2017 Anderson et al. |
| [79] 2017 Chiu et al. | [80] 2017 Johnson et al. |
| [81] 2017 Stuart et al. | [82] 2017 McGillen et al. |
| [83] 2017 Cremin et al. | [84] 2018 Ross et al. |
| [85] 2018 Anderson et al. | [86] 2018 Anderson et al. |
| [87] 2018 Omondi, Mbogo, and Luboobi | [88] 2018 Woods et al. |
| [89] 2018 Stevens et al. | [90] 2019 Stopard et al. |
| [91] 2019 Beacroft and Hallett | [92] 2019 Reidy et al. |
| [93] 2019 Omondi, Mbogo, and Luboobi | [94] 2019 Maheu-Giroux et al. |

## B Definitions & Extraction

Data were obtained from (in order of precedence): article text; article tables; article figures; appendix text; appendix tables; appendix figures; and likewise for articles cited like “the model is previously described elsewhere”. Data were assessed from figures with the help of a graphical measurement tool.^1^

**Fitted Parameters:** For the values of fitted parameters, we used the posterior value as reported, including the mean or median of the posterior distribution, or the best fitting value. If the posterior was not reported, we used the mean or median of the prior distribution, including the midpoint of uniform sampling ranges.

### B.1 Epidemic Context

Let *t_0_* be the time of ART scale-up/scenario divergence in the model.

**HIV Prevalence:** As reported in the context overall at *t_0_*: *Low*: *<*1%; *Medium*: 1-10%; *High*: *>*10%.

**Epidemic Phase:** As projected in the base-case scenario in the context overall between *t_0_* and roughly *t_0_* + 10 years: *Increasing* (linear or exponential); *Increasing but stabilizing*; *Stable*; *Decreasing but stabilizing*; *Decreasing* (linear or exponential).

**Geographic Scale:** For studies of one geographic context, scale was defined as one of: *regional*: multiple countries; *national*: one country; *sub-national*: smaller than a country but greater than a city; *city*: one city or less. For studies that consider multiple geographic contexts, scale was defined as *multi-x*, where *x* is the smallest geographically homogeneous scale considered from the list above.

**Country:** The countries counted were: Angola, Benin, Botswana, Burkina Faso, Burundi, Cameroon, Cape Verde, Central African Republic, Chad, Comoros, Democratic Republic of the Congo, Djibouti, Equatorial Guinea, Eritrea, eSwatini, Ethiopia, The Gabon, Gambia, Ghana, Guinea, Guinea-Bissau, Côte d’Ivoire, Kenya, Lesotho, Liberia, Madagascar, Malawi, Mali, Mauritania, Mauritius, Mozambique, Namibia, Niger, Nigeria, Republic of the Congo, Reunion, Rwanda, Sao Tome and Principe, Senegal, Seychelles, Sierra Leone, Somalia, South Africa, South Sudan, Sudan, Tanzania, Togo, Uganda, Zambia, Zimbabwe. See Table A.1 for related search terms. If a study modelled multiple countries at a national scale or smaller, the counter for each country was incremented.

### B.2 Risk Heterogeneity

#### B.2.1 Key Populations

**Female Sex Workers:** Any female activity group meeting 3 criteria: representing *<*5% of the female population; and being *<*1/3 × the size of client population or highest non-MSM male activity group; and having *>*50 × the partners of the lowest sexually active female activity group [95, 96, 97]. We also noted whether the authors described any activity groups as FSW. If it was not possible to evaluate any criteria due to lack of data, then we assumed the criteria was satisfied.

**Clients of FSW:** Any male activity group meeting 2 criteria: described as representing clients of FSW; being *>*3 × the size of the FSW population [96]. If group sizes were not reported, then we assumed an activity group described as clients met the size criterion. We also noted whether clients were described as comprising a proportion of another male activity group.

**Men who have Sex with Men:** Any male activity group(s) described by the authors as MSM.

**Transgender People:** Any activity group(s) described by the authors as transgender.

**People who Inject Drugs:** Any activity group(s) described by the authors as PWID.

**Prisoners:** Any activity group(s) described by the authors as prisoners.

#### B.2.2 Activity Groups

Activity groups were defined as any stratification based on sex/gender and the number and/or types of partnerships formed, including key populations, but excluding stratifications by age.

**Count:** We counted the number of modelled activity groups in total, and separately for women who have sex with men, men who have sex with women, and MSM.

**Highest Risk Group Size:** The proportion of men and women in the highest risk group.

**Turnover:** Turnover refers to movement of individuals between activity groups and/or key populations reflecting sexual life course. We defined four classifications of turnover if activity groups were modelled: *None*: no movement between activity groups; *High-Activity*: only movement between one high activity group or key population and other activity group(s); *Multiple*: movement between multiple pairs of risk groups; *Replacement*: only movement from low to high activity to maintain high activity group size(s) against disproportionate HIV mortality.

#### B.2.3 Partnerships

**Approaches:** How studies defined partnerships, classified into one of three approaches: *Generic*: all partnerships are equal; *By-Group:* partnership types are defined only by the activity groups involved; *Overlapping:* multiple partnership types can be formed by the same pair of activity groups. Within *By-Group*, we classified how the parameters of the partnership were defined, as based on either: the *susceptible* partner; the *lower activity* partner; the *higher activity* partner; or some consideration of *both partners*.

**Characteristics:** Whether any of the following varied between different partnership types: *Condom Use*: proportion of sex acts protected; *Total Sex*: total number of sex acts, possibly defined by differences in partnership duration and/or frequency of sex.

**Mixing:** Mixing by activity group was classified as either: *Proportionate*: proportionate to the total number of partnerships offered by each risk group; *Assortative*: any degree of preferential partnership formation between individuals of the same or similar risk groups.

#### B.2.4 Age Groups

**Count:** The number of age groups considered in the model.

**Risk:** Whether age groups differed in any characteristic that conferred transmission risk (binary).

**Mixing:** We classified whether partnership formation between age groups was assumed to be: *Proportionate*: proportionate to the number of partnerships offered by each age group; *Strictly Assortative*: any degree of preferential partnership formation between individuals of the same or similar age groups that is equal for both sexes. *Off-Diagonal*: any degree of preferential partnership formation between younger women and older men.

### B.3 HIV Natural History

**Count:** The number of states of HIV infection considered in the model, excluding stratifications related to treatment. If states were defined by both CD4 and viral load, then the count considers all unique combinations.

**Acute Infection:** Whether any state represented increased infectiousness associated with acute infection (binary).

**Late-Stage Infection:** Whether any state(s) considered increased infectiousness associated with late-stage infection (binary).

**HIV Morbidity:** Whether any state(s) considered decreased sexual activity associated with late-stage disease (binary), and how that decreased was modelled: *Inactive*: complete cessation of sexual activity; *Partners*: decreased rate of partnership formation; *Sex Acts per Partnership*: decreased frequency of sex per partnership; and/or *Generic*: representative decreased probability of transmission.

### B.4 Antiretroviral Therapy

#### B.4.1 Transmission

**Transmission Reduction due to ART:** The relative reduction in probability of transmission (0 is perfect prevention, 1 is no effect) among individuals who are virally suppressed; if viral suppression was not explicitly modelled, then the relative reduction among individuals who are on treatment was used.

**Transmitted Resistance:** Any consideration of 1+ strains of HIV which are transmitted and for which ART had reduced benefits. We did not document the number of resistant strains, or characteristics of resistance and transmissibility.

#### B.4.2 Treatment Cascade States

**Forward Cascade:** We extracted whether each of the following states were included (binary): *Diagnosed*: aware of their HIV+ status, but have not yet started ART; *Not Yet Virally Suppressed*: started ART, but are not yet virally suppressed; *Virally Suppressed*: on ART and achieved viral suppression; and *Generic On ART* : simplifications of any/all of the above.

**Stopping ART:** We extracted whether individuals stopped ART, either due to: *Treatment Failure*: ART is no longer efficacious due to resistance; or *ART Cessation*: ART is discontinued for other reasons, such as barriers to access or side effects. We also extracted whether individuals stopping ART for either reason were tracked separately, or whether they re-entered a generic ART-naive state, such as “Diagnosed”.

**Differential Cascade Transitions:** We extracted whether rates of transitioning along the ART cascade, including: rate of *HIV diagnosis*; rate of *ART initiation*; and rate of *ART cessation*, differed by any of the following stratifications: *sex*; *age*; *activity*; and *key populations*. If the study did not mention possible differences in such rates, then we assumed that no differences were modelled.

#### B.4.3 Behaviour Change

**HIV Counselling:** Whether any sexual behaviour change associated with HIV testing and counselling was applied to individuals in the diagnosed and/or on-ART states (binary), and what changed: *Condom Use*: increased; *Serosorting*: any; *Partners*: decreased rate of partnership formation; *Sex Acts per Partnership*: decreased frequency of sex per partnership; and/or *Generic*: representative decreased probability of transmission due to counselling.

### B.5 ART Prevention Impact

The following data were extracted per scenario, rather than per-study.

#### B.5.1 Intervention

**ART Initiation Criteria:** What criteria were used for ART eligibility as part of the intervention: *Symptomatic (AIDS)*; *CD4 < 200*; *CD4 < 350*; *CD4 < 500*; *All individuals*; *Other*.

**Intervention Population:** Among which population sub-group(s) was the scale-up of ART coverage/initiation applied. Only scenarios with ART intervention for all individuals were included in Dataset B.

**Impact Population:** Among which population sub-group(s) was the ART prevention impact measured. Only scenarios measuring ART prevention impacts in all individuals were included in Dataset B.

**ART Coverage Target:** The proportion of people living with HIV in the intervention population who are on ART by the end of ART scale-up.

**ART Initiation Rate Target:** The rate at which people living with HIV in the intervention population initiate ART by the end of ART scale-up.

**Intervention *t_0_* and *t_f_*:** The years at which ART scale-up as part of the intervention started and stopped, respectively. If interventions were modelled as instantaneous, such as increasing ART initiation rate, then we considered *t_0_* = *t_f_*. Impact time horizons were measured relative to *t_0_*.

#### B.5.2 Impact

For both measures of ART prevention impact, we extracted reported values from the text for any available time horizon, as well as figure data for any of the following time horizons, if available: 5, 10, 15, 20, 30, and 40 years, with the help of a graphical measurement tool. If only absolute values were reported, we calculated the relative reductions manually. Where reported, we extracted confidence intervals for each outcome.

**Relative Incidence Reduction:** The relative reduction in overall annual HIV incidence (per 1000 person-years) in the intervention scenario as compared to the baseline scenario, both after an equal number of years since *t_0_* (time horizon). For example, if the baseline and intervention scenarios predicted overall HIV incidence of 1 and 0.7 per 1000 person-years 5 years after *t_0_*, then the relative incidence reduction for the 5-year time horizon would be 30%.

**Proportion of Infections Averted:** The relative reduction in cumulative new HIV infections in the intervention scenario as compared to the baseline scenario, both after an equal number of years since *t_0_* (time horizon). For example, if the baseline and intervention scenarios predicted 1000 and 700 new infections 5 years after *t_0_*, then the proportion of infections averted for the 5-year time horizon would be 30%.

## C Supplemental Results

Additional information on data sources, analysis, and results are available in the public repository:

https://github.com/mishra-lab/sr-heterogeneity-hiv-models

### C.1 Map

**Figure C.1:**
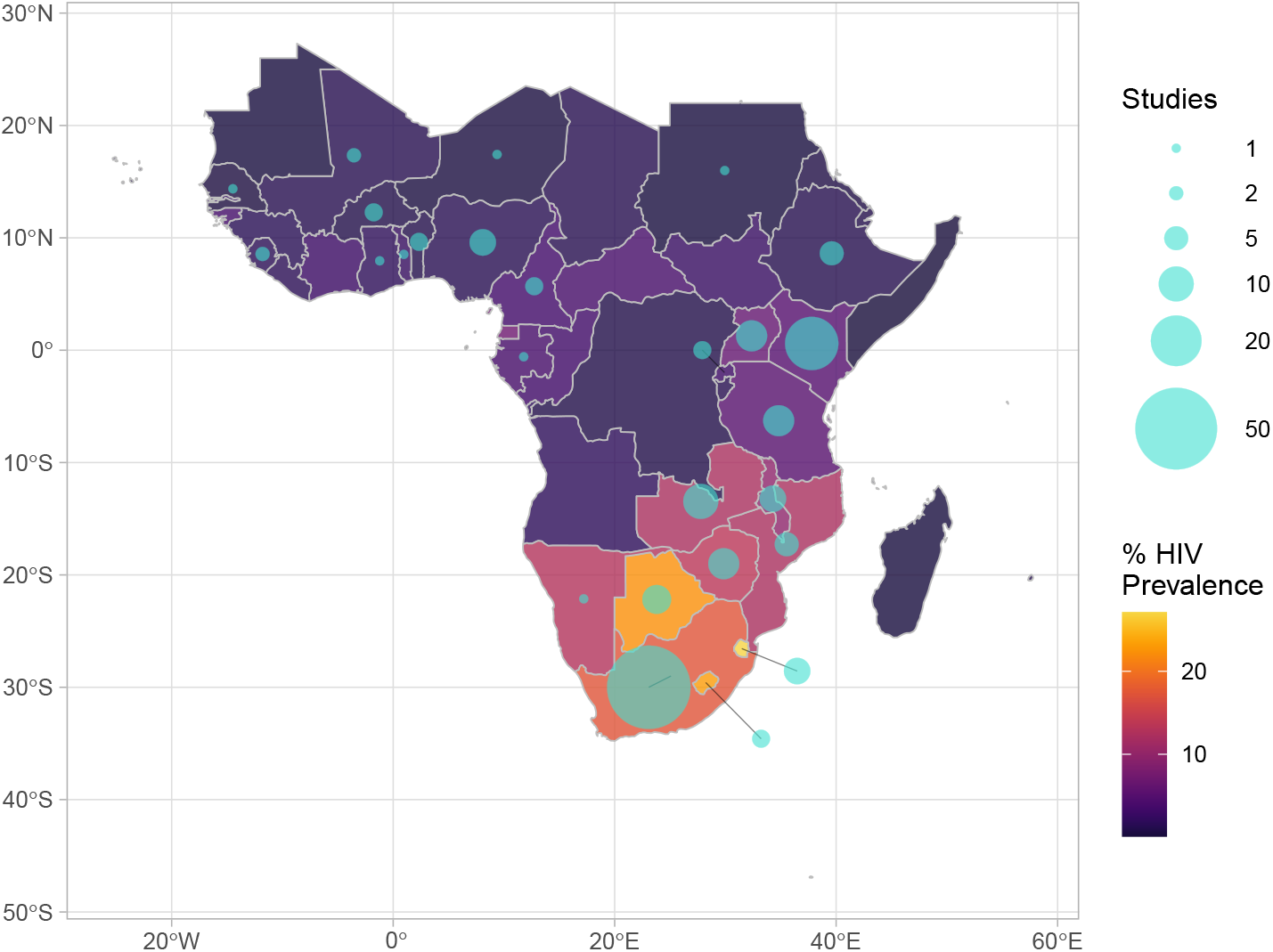
Map showing number of studies (of 94 total) applying HIV transmission modelling in each country vs the number of people living with HIV (PLHIV, millions)

### C.2 Risk Heterogeneity

#### C.2.1 Distributions

The following figures illustrate the distributions (number of studies) of various parameter values and modelling assumptions related to factors of heterogeneity and intervention contexts.

**Figure C.2:**
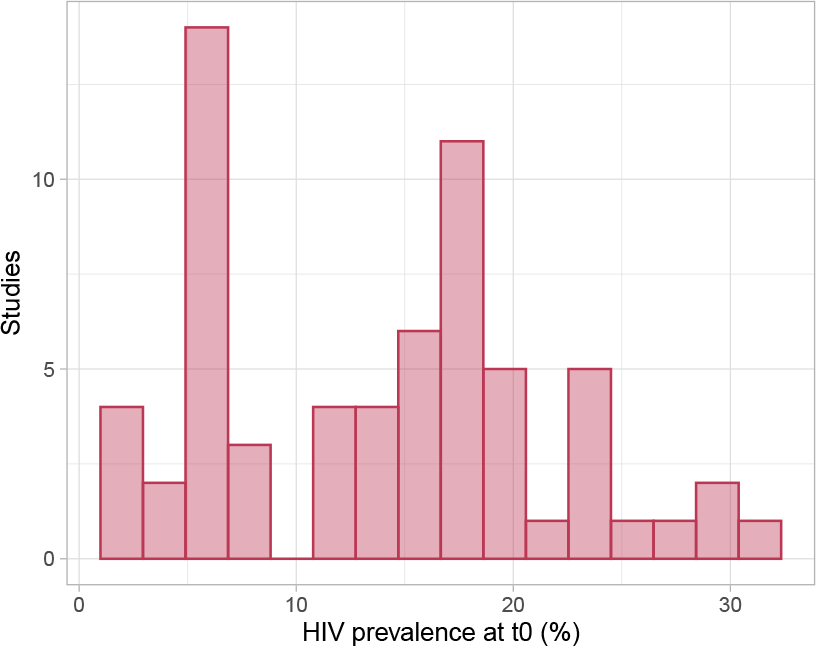
HIV prevalence at **t**_0_ (%)

**Figure C.3:**
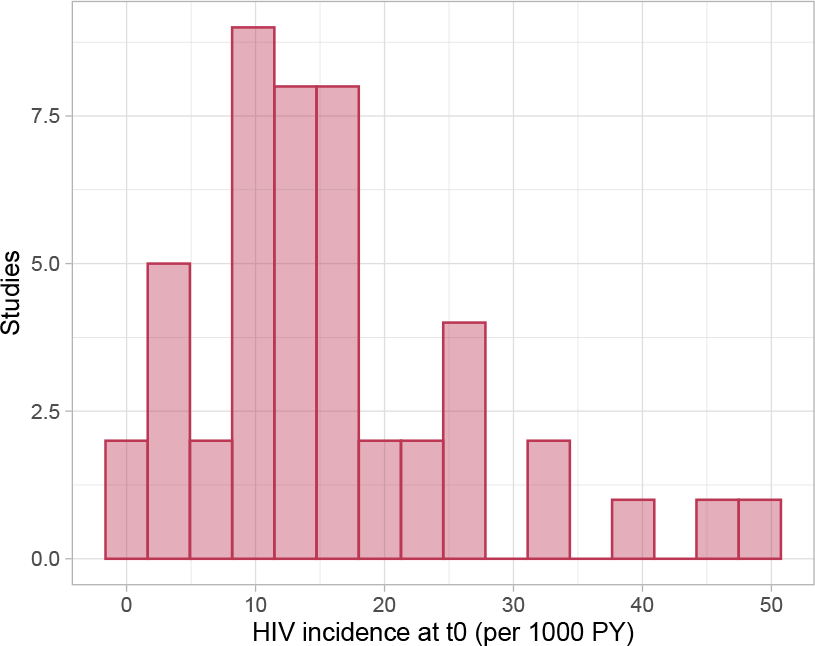
HIV incidence at **t**_0_ (per 1000 PY)

**Figure C.4:**
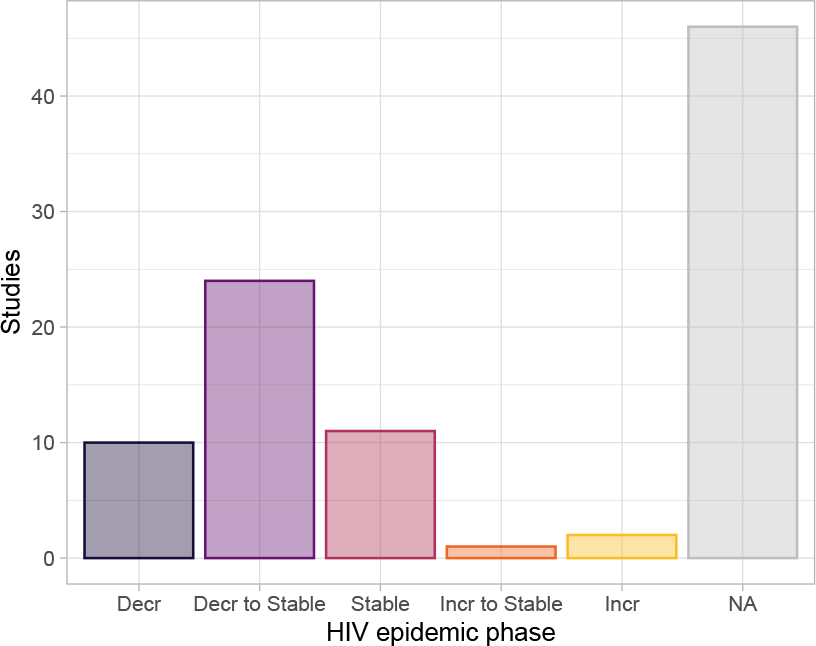
HIV epidemic phase

**Figure C.5:**
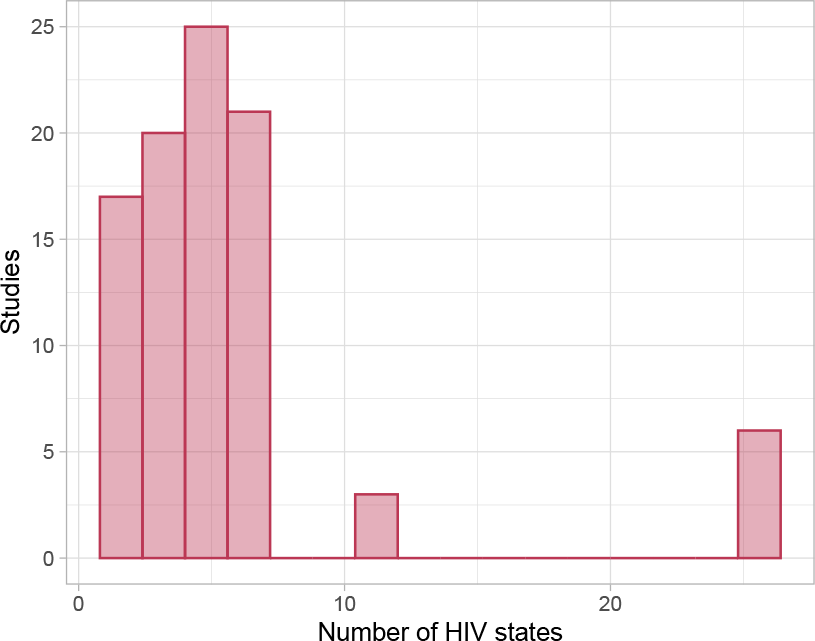
Number of HIV states

**Figure C.6:**
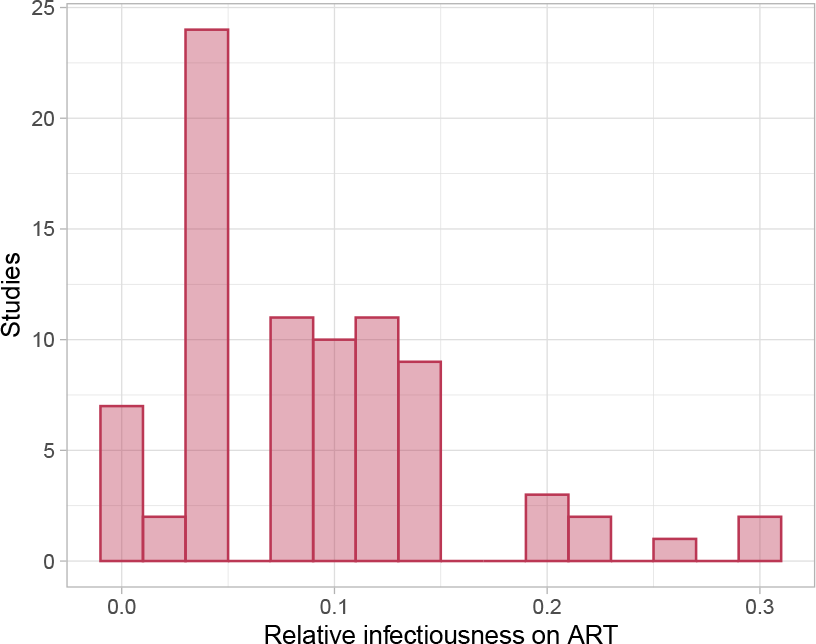
Relative infectiousness on ART

**Figure C.7:**
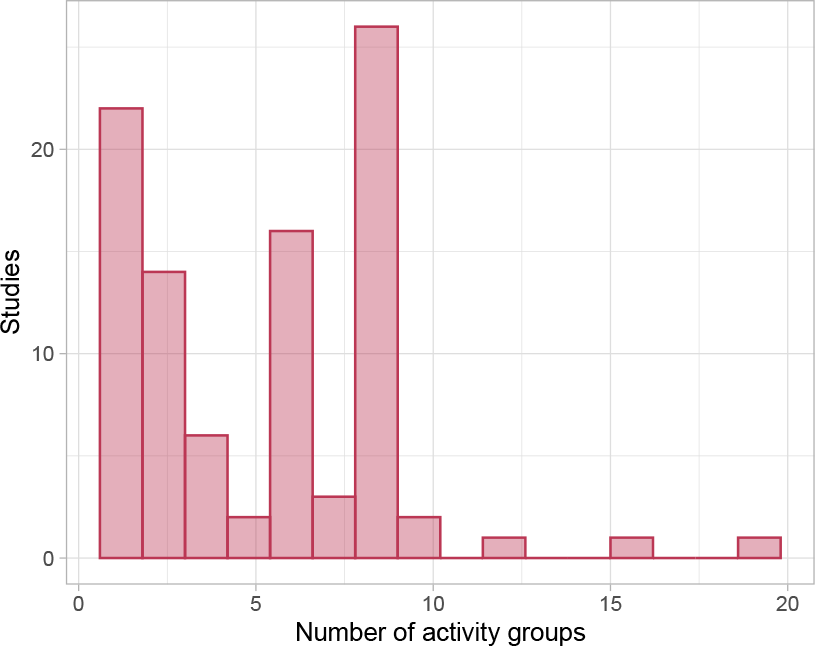
Number of activity groups

**Figure C.8:**
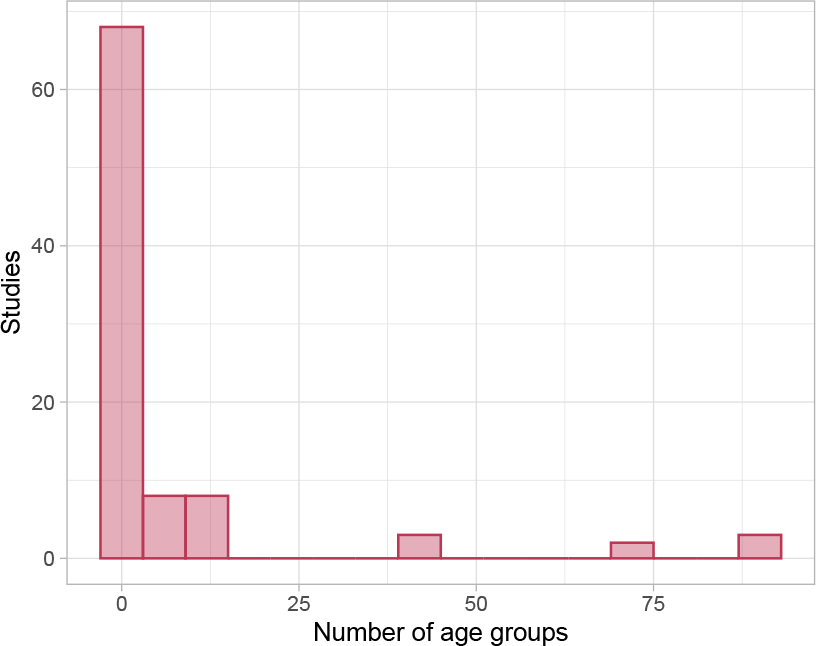
Number of age groups

**Figure C.9:**
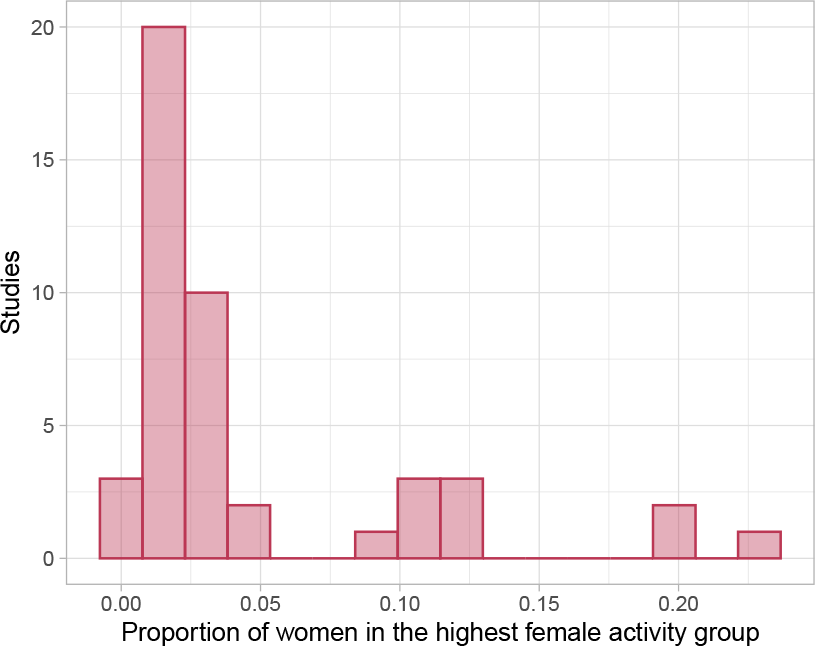
Proportion of women in the highest female activity group

**Figure C.10:**
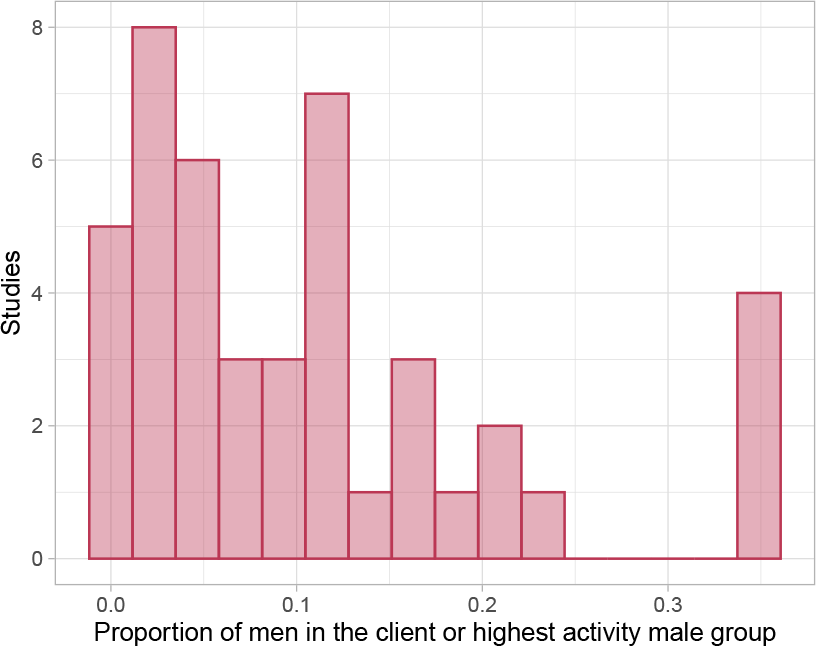
Proportion of men in the client or highest activity male group

### C.3 ART Prevention Impact

The following figures illustrate the projected ART prevention impact (Dataset B), stratified by various factors of heterogeneity and intervention contexts (colours). Left panels show the relative HIV incidence reduction (IR); right panels show the proportion of cumulative HIV infections averted (CIA); both as compared to a base-case scenario reflecting status quo. If any study included multiple scenarios of ART scale-up, then each scenario was included separately; if any scenario reported multiple time horizons, each time horizon was included separately. The number of studies (scenarios) reporting incidence reduction, cumulative infections averted, both, or either was: 23 (61), 24 (75), 7 (11), and 40 (125), respectively. If any factor could not be quantified due to missing data or varying values, it was omitted from that plot. In box plots, the numbers of unique scenario time-horizons contributing to each box are given above it.

**Figure C.11:**
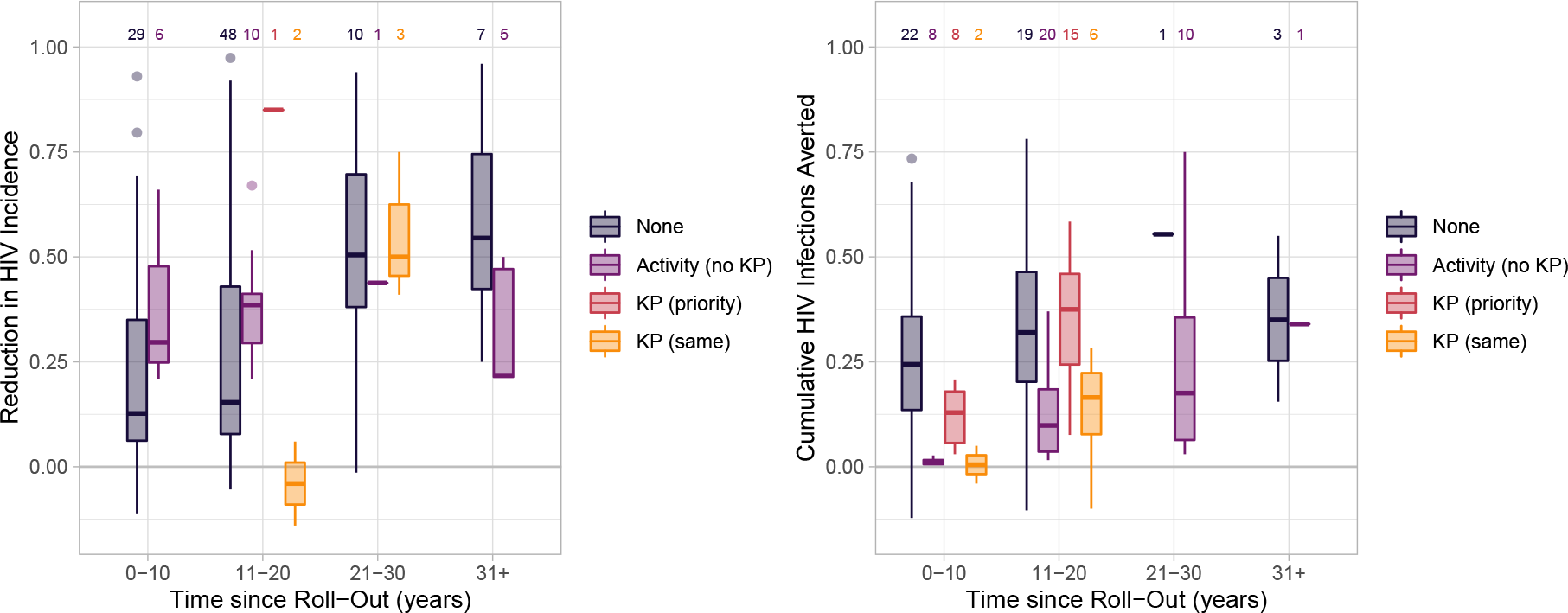
Risk Stratification & ART cascade differences

**Figure C.12:**
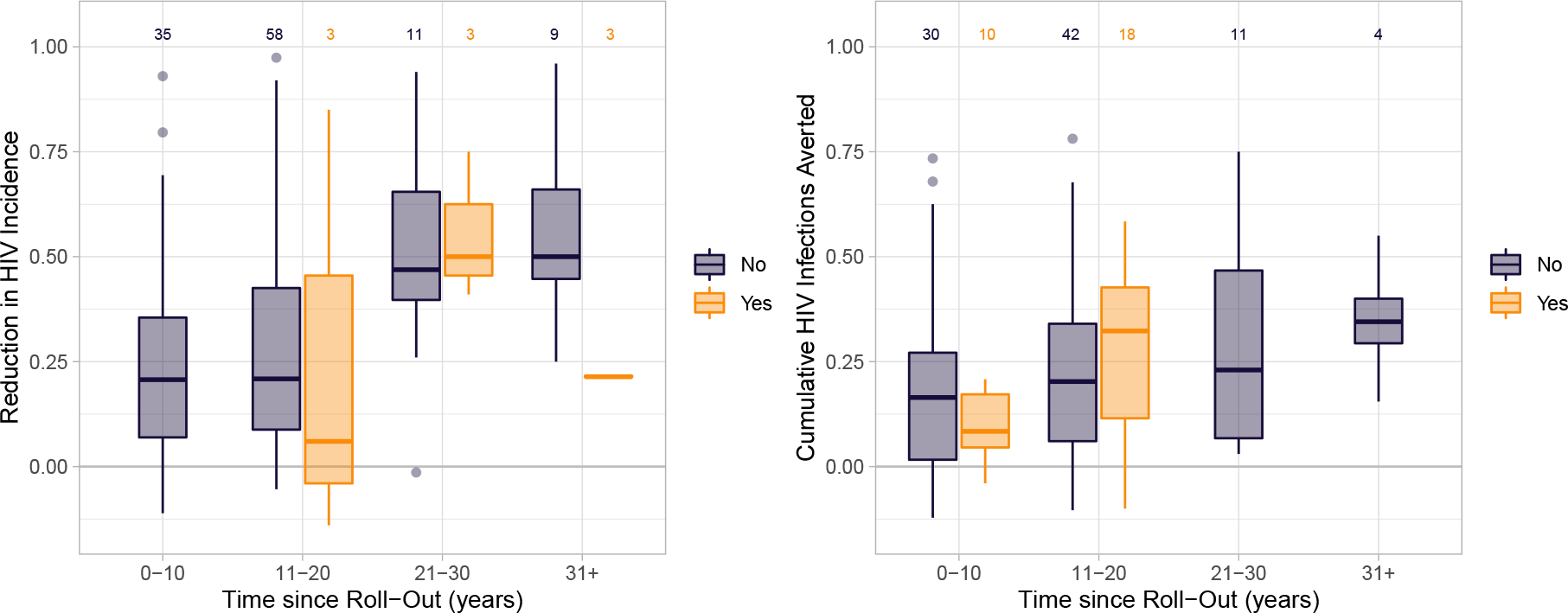
Any activity group turnover

**Figure C.13:**
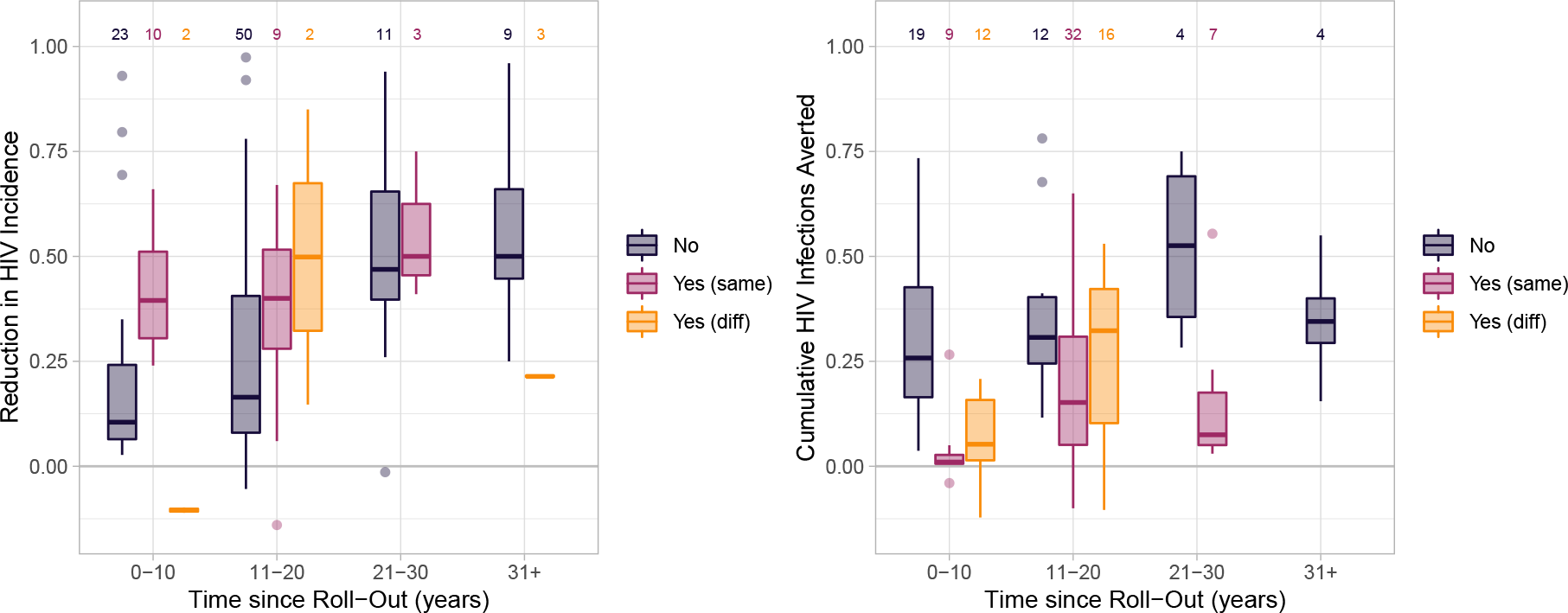
Sex stratification & any ART cascade differences

**Figure C.14:**
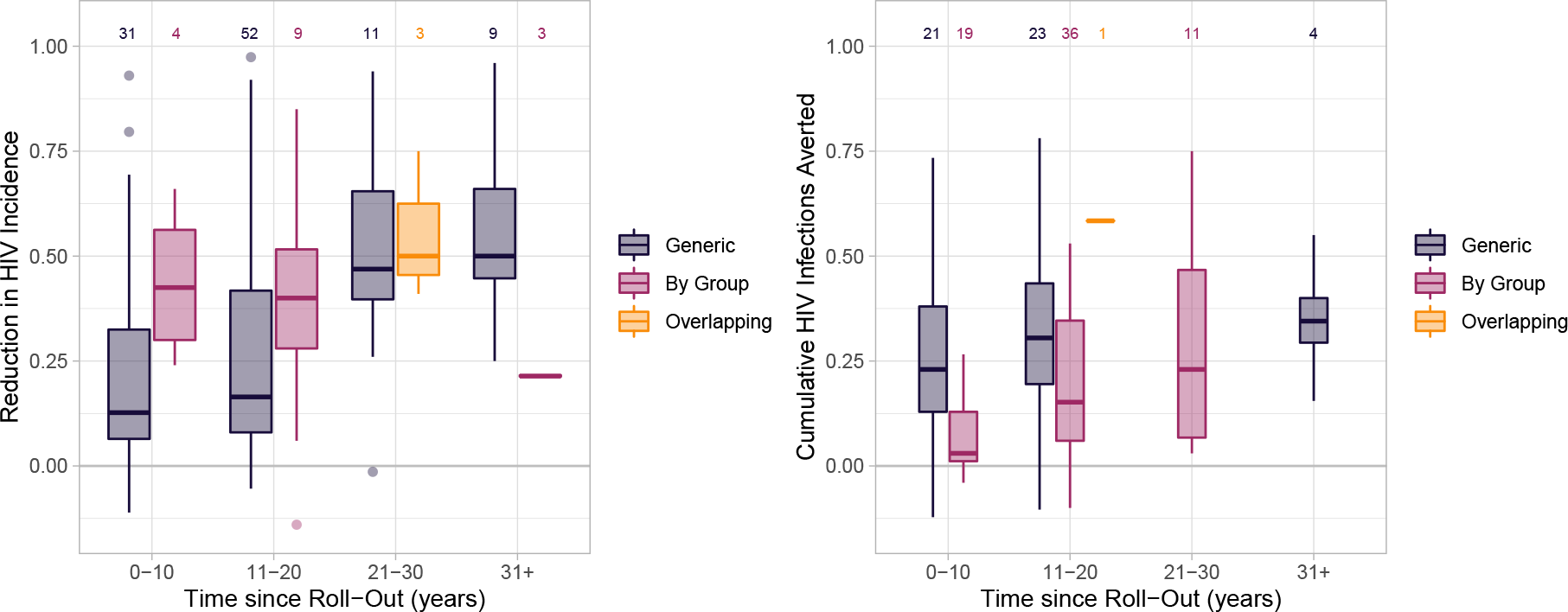
Type of partnership definition

**Figure C.15:**
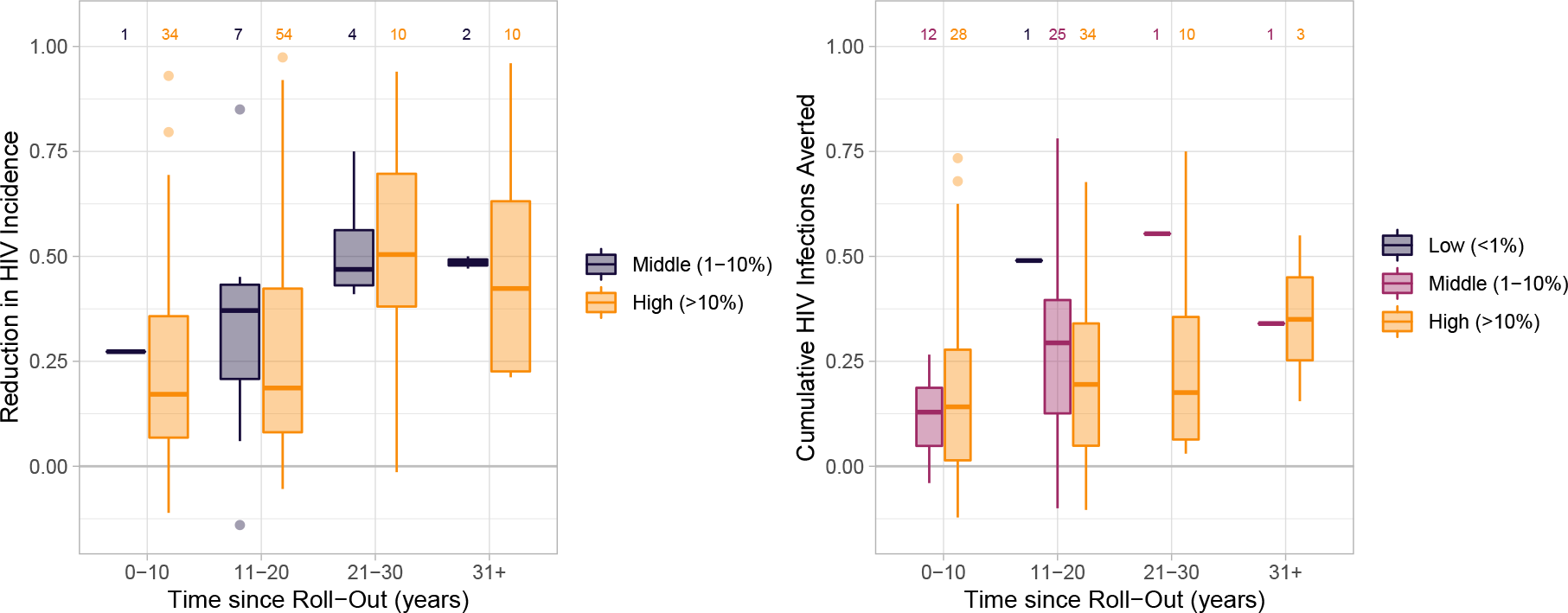
HIV prevalence at **t**_0_ (%)

**Figure C.16:**
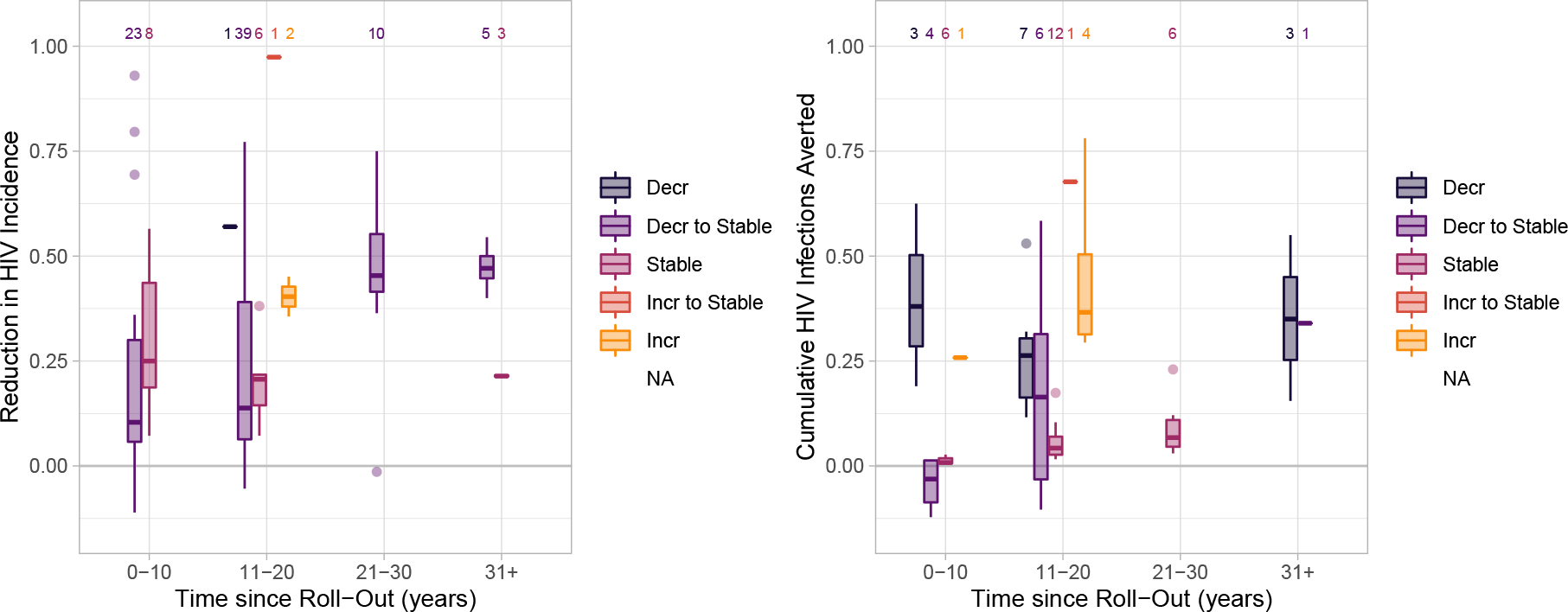
HIV epidemic phase

**Figure C.17:**
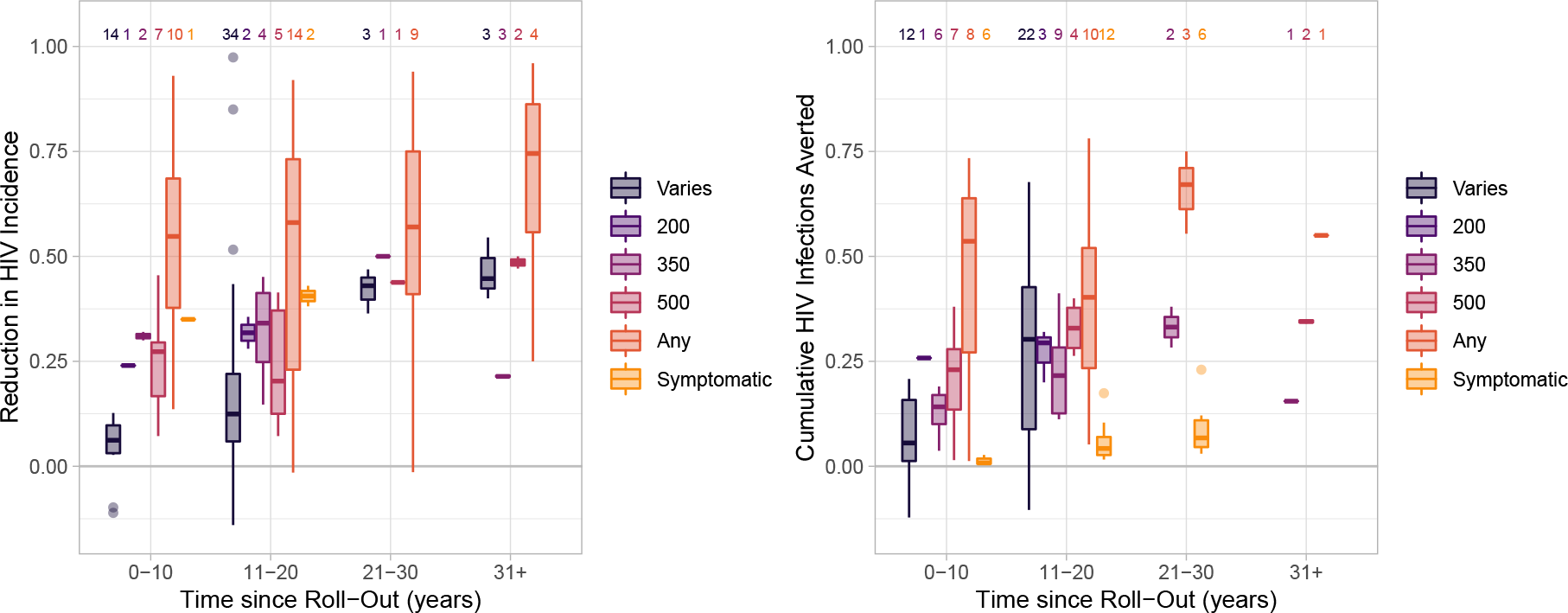
CD_4_ initiation criteria

**Figure C.18:**
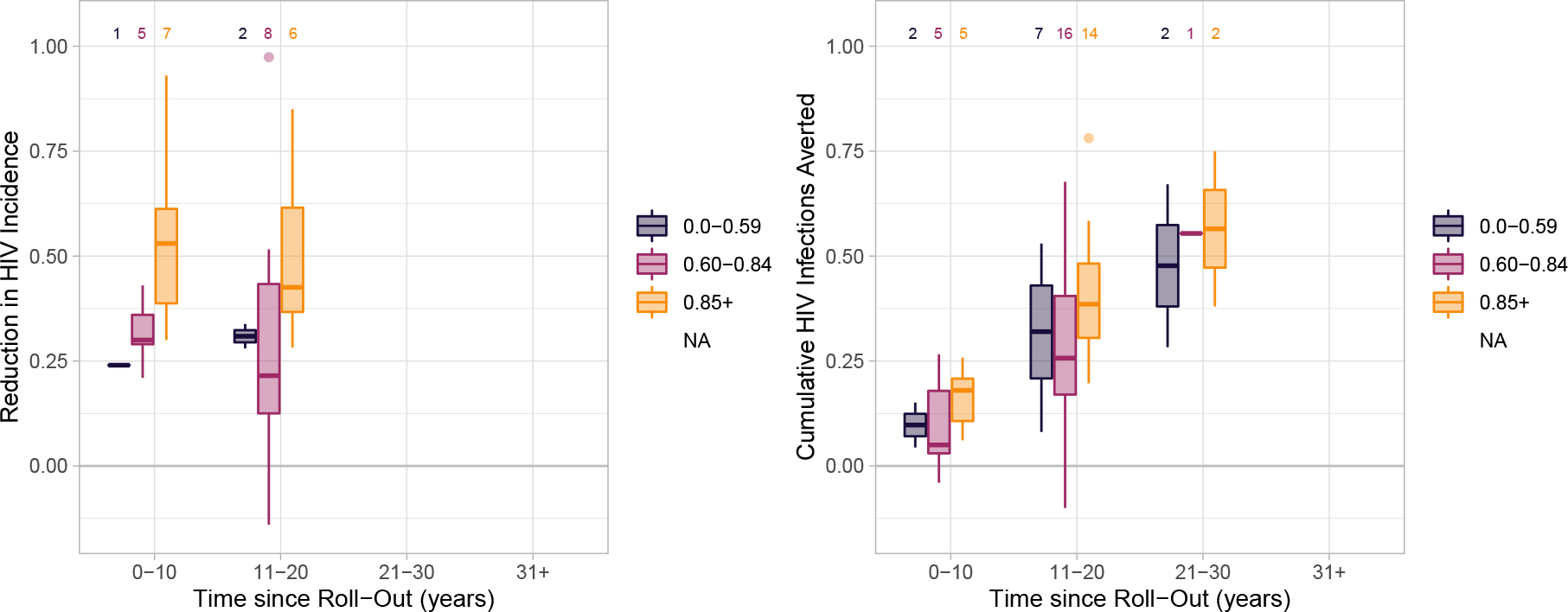
ART intervention coverage target

**Figure C.19:**
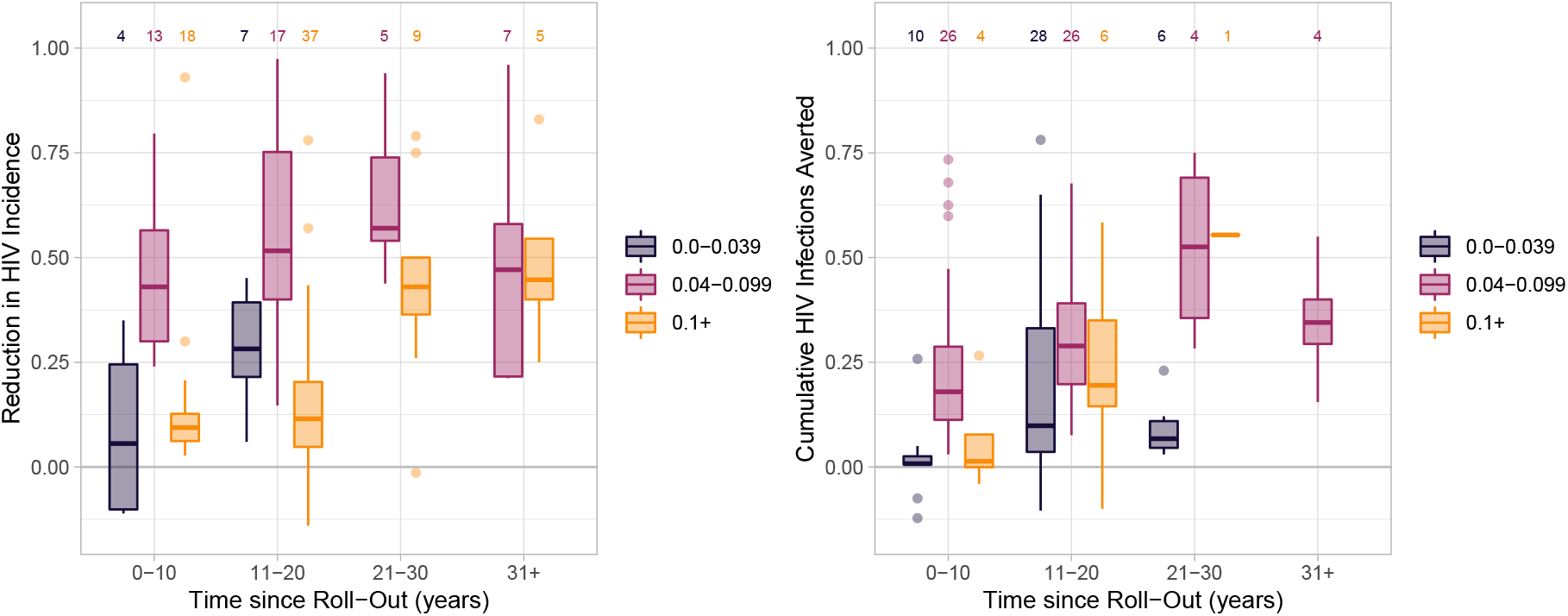
Relative infectiousness on ART

**Figure C.20:**
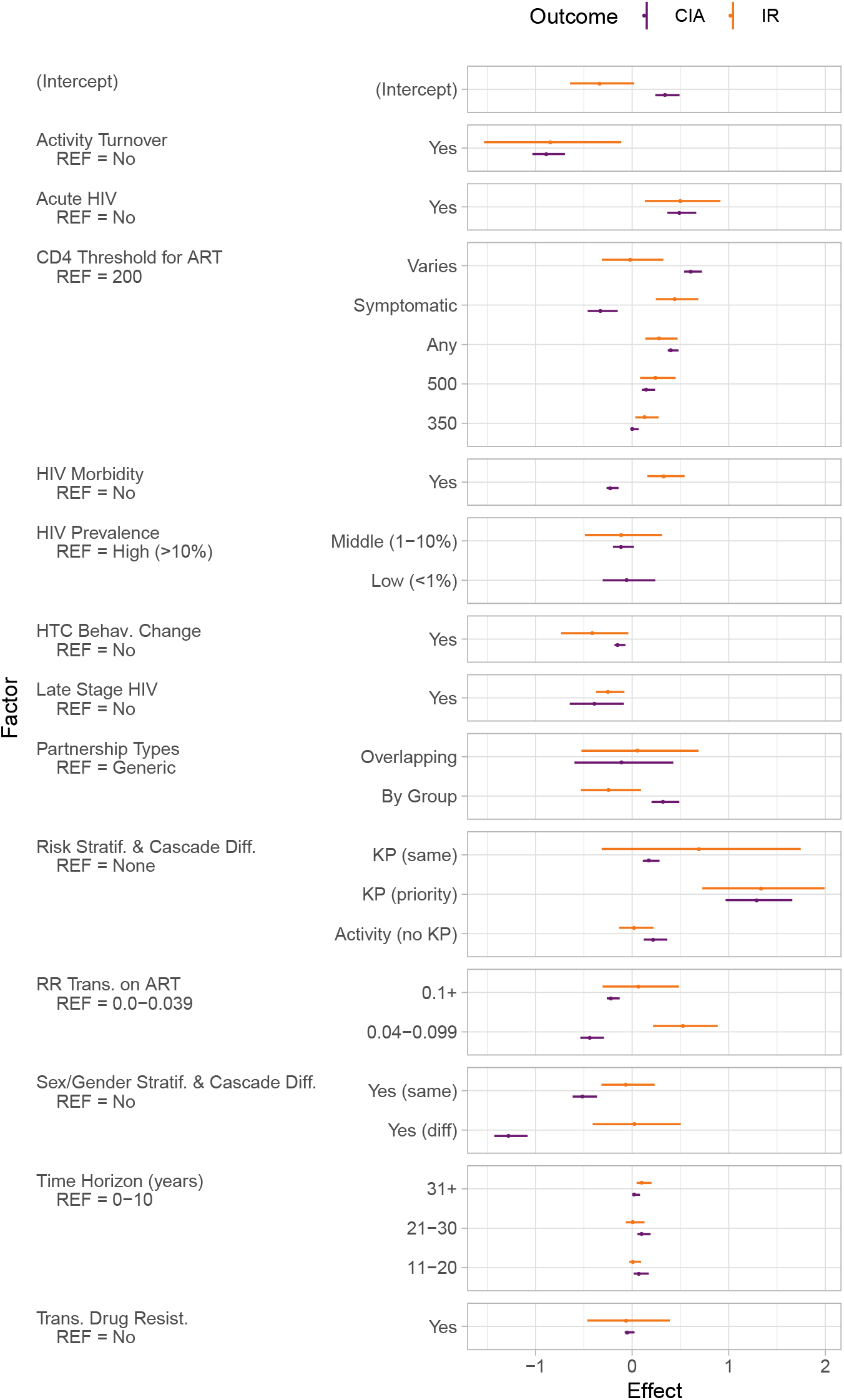
Effect estimates for factors of heterogeneity on incidence reduction (%, IR) and cumulative infections averted (%, CHI) from linear multivariate regression with generalized estimating equations. Numerical results given in Table 3. RR: relative risk; HTC: HIV testing and counselling; KP: key populations. priority: modelled ART cascade transitions were faster in KP vs overall due to prioritized programs; same: cascade transitions were assumed the same in KP as overall. Factor definitions are given in Appendix B.

## D PRISMA-ScR Checklist

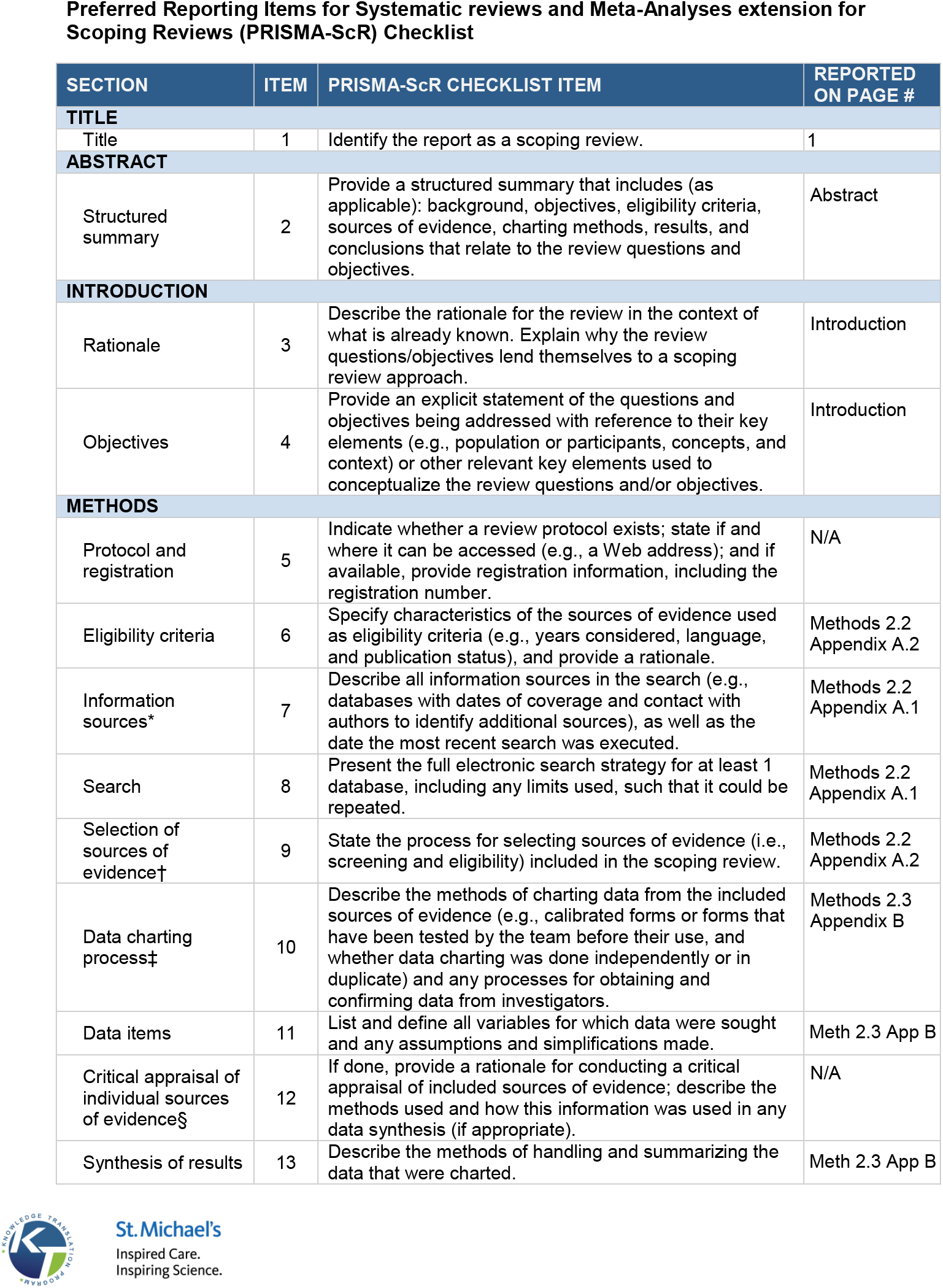
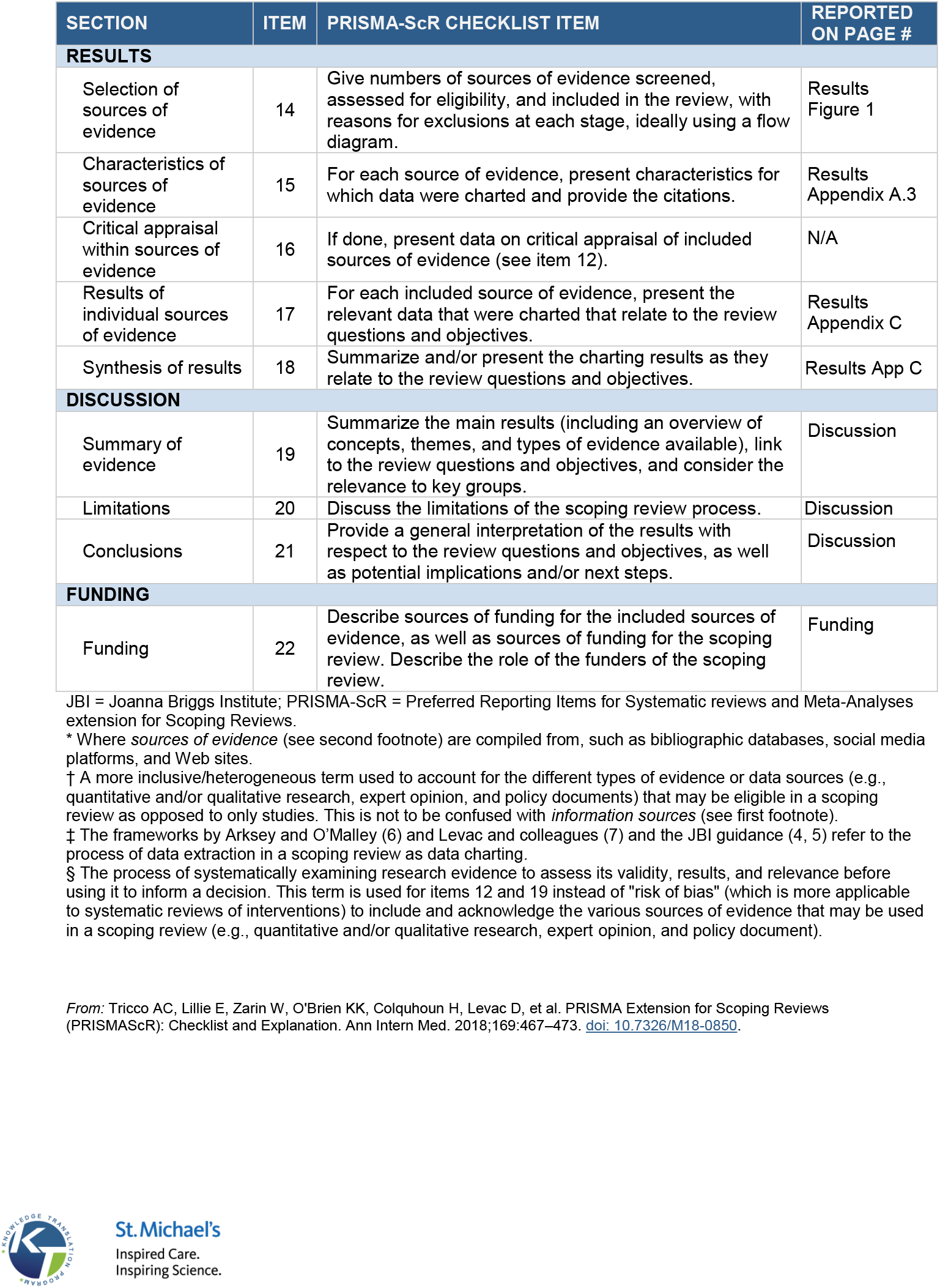

